# SPARC: A mechanism-aware spatial representation from routine histology predicts cancer survival and therapy response

**DOI:** 10.64898/2026.05.04.26352410

**Authors:** Aziz Ayed, Gabrielle Cohn, Noé Bertramo, Genevieve Boland, Justin Gainor, Ömer H. Yilmaz, Regina Barzilay

## Abstract

Understanding the molecular mechanisms that drive treatment response is central to personalized cancer care, but assays such as spatial transcriptomics are not yet scalable in routine clinical practice. A critical question, then, is whether this deeper molecular insight can be extracted directly from routine histology. Here, we introduce SPARC, a framework that infers spatially resolved activity maps for 40 gene expression programs directly from H&E slides. Integrating predicted program maps with morphological features improves survival prediction in 17 of 18 cancer types across 8,383 patients and matches a multi-omic method requiring paired RNA sequencing. SPARC also stratifies bevacizumab response in ovarian cancer (odds ratio = 8.08) and trastuzumab response in breast cancer (odds ratio = 3.44), while H&E image-only baselines yield non-significant separation between responders and non-responders. Unsupervised analysis of predicted maps reveals canonical tumor microenvironment compartments and spatial interaction patterns directly from tissue morphology, linking predictive performance of clinical outcomes to underlying biological mechanisms.

## Introduction

Understanding the molecular mechanisms that drive tumor development and treatment response is central to personalizing cancer care [1]. Comprehensive molecular assays such as spatial transcriptomics and multiplexed immunofluorescence are able to shed light on these mechanisms by characterizing the complex biology of the tumor microenvironment. However, these technologies are not used in routine clinical practice due to cost and scalability limitations [2, 3]. In contrast, hematoxylin and eosin (H&E)-stained pathology slides are ubiquitously collected for diagnostic purposes across virtually every cancer diagnosis and care pathway [4]. Beyond their established role in diagnosis, however, these slides contain a wealth of morphological information that reflects the underlying biology of the tumor. A critical question, then, is whether deeper molecular insights can be extracted directly from routine H&E slides, without requiring specialized molecular assays. Here, we show that this is possible by modeling gene expression programs—coordinated sets of genes that reflect defined biological processes [5, 6]—and inferring their activity from tissue morphology.

Programs provide an interpretable and clinically actionable view of tumor biology. As one example of a program, consider angiogenesis, the process by which tumors recruit new blood vessels to sustain growth. The activity of this program serves as a predictor of both patient prognosis and the efficacy of therapies such as bevacizumab in colorectal cancer [7]. In this work, angiogenesis is one of 40 biological programs we learn to predict, drawn from pathway databases and literature-derived signatures to collectively span the major axes of tumor biology, from proliferation and metabolism to immune signaling and microenvironmental remodeling. Critically, such programs also leave identifiable visual signatures in tissue morphology—angiogenesis remodels the tumor vasculature in ways that are visible on H&E-stained sections as aberrant vessel architecture and altered stromal organization [8]. This morphological footprint suggests that a computational model can learn to infer program activity from tissue appearance alone.

Beyond interpretability, program-level representations provide a biologically grounded signal for downstream modeling. Existing computational pathology approaches primarily indicate which tissue regions drive the model’s predictions [9], but do not capture the biological basis of these associations. In contrast, program-level predictions link spatial patterns to defined processes such as immune exclusion or metabolic reprogramming, thereby clarifying why specific regions may be clinically relevant. By distilling complex histologic variation into a structured set of programs, this representation also reduces the dimensionality of the learning task, yielding models that are more stable, require fewer samples to train, and achieve improved prognostic performance in the small-cohort settings typical of clinical studies.

Prior computational approaches have addressed parts of this problem but face important limitations. Virtual staining and spatial transcriptomics prediction methods attempt to reconstruct individual gene expression values from H&E [10], but predicting thousands of genes at spatial resolution remains extremely challenging, often yielding limited performance. In contrast, modeling coordinated biological programs provides a more stable and biologically meaningful target. Separately, multi-omic integration methods have demonstrated that combining histological features with molecular data can improve prognostic accuracy [11], but these approaches require paired molecular profiling—such as bulk RNA sequencing—at inference time, limiting their scalability and clinical applicability. We therefore bridge these two lines of work by introducing SPARC (Spatial Prediction of Activity and Response in Cancer), a framework that infers spatially resolved activity maps for 40 gene expression programs directly from H&E slides and integrates these biological representations with morphological features for clinical prediction, requiring only a standard H&E slide as input.

We evaluated SPARC across multiple clinical settings. Unsupervised analysis of predicted program maps recovered canonical tumor microenvironment compartments and spatial interaction patterns directly from tissue morphology. SPARC-Risk improved survival discrimination in 17 of 18 cancer types across The Cancer Genome Atlas and four independent cohorts, performing comparably to a multi-omic method that requires paired RNA sequencing [11]. In two clinical cohorts, SPARC achieved significant stratification of bevacizumab response in ovarian cancer and trastuzumab response in breast cancer—including in small cohorts of 73 and 85 patients, respectively—with predictive programs aligning closely with independently reported molecular subtypes of treatment response [12].

## Results

### Study overview

SPARC is a two-stage computational framework that maps routine histopathology to spatially resolved biological programs and links these representations to clinical outcomes (Fig. 1). In the first stage, SPARC-Map learns to identify where our 40 curated gene expression programs are active within a tumor using standard H&E-stained tissue slides. To generate training annotations for learning these programs, we used spatial transcriptomics data to estimate the activity of these processes at each slide patch. Specifically, gene expression measurements were converted into program-level scores using ssGSEA. In the second stage, SPARC-Risk combines these predicted maps of biological activity with detailed image features extracted by a large pretrained pathology model (H-Optimus-1) [13], and uses this information to generate patient-level risk predictions. We trained SPARC-Risk on the pan-cancer TCGA cohort [14] to predict disease-specific survival (DSS) and evaluated the resulting model on external datasets in both zero-shot and fine-tuned settings across multiple clinical endpoints. We further assessed whether the predicted program maps capture treatment-relevant biology by testing their ability to stratify response to select anti-cancer therapies, including bevacizumab in ovarian cancer and trastuzumab in breast cancer. Finally, we leveraged the spatial structure of predicted program activity to define tumor microenvironment (TME) niches, enabling a higher-order characterization of tissue organization and its association with clinical outcomes.

**Figure 1:**
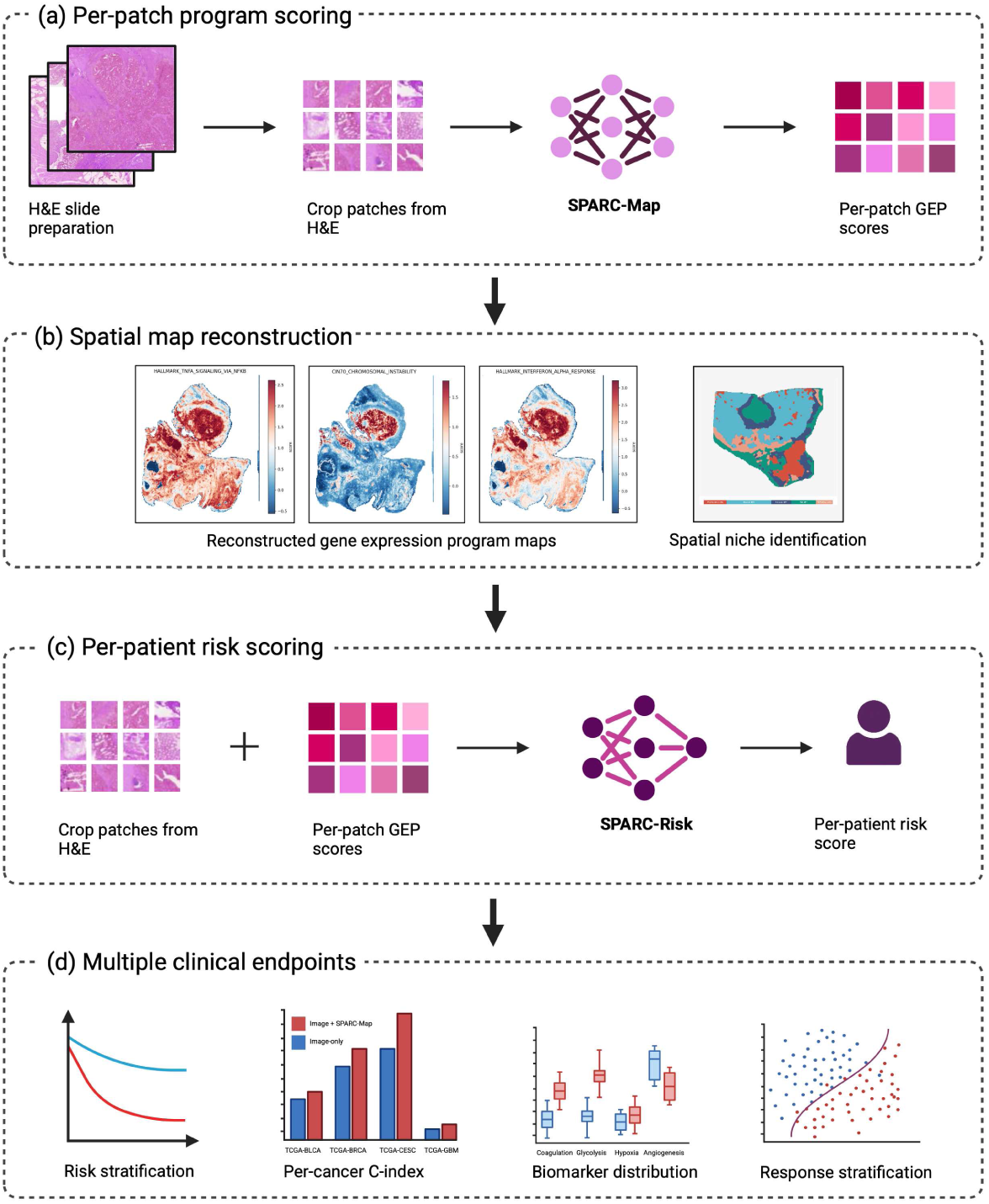
Overview of the SPARC framework for inferring spatial gene expression programs and predicting clinical outcomes from H&E slides. **(a)** Whole-slide H&E images are tiled into patches and processed by SPARC-Map to predict patch-level gene expression program (GEP) scores learned from paired spatial transcriptomics data. **(b)** Patch-level predictions are aggregated to reconstruct spatial maps of GEP activity, enabling identification of intratumoral heterogeneity and spatial niches. **(c)** Patch-level image features and predicted GEP scores are integrated by SPARC-Risk to derive patient-level risk scores. (d) SPARC-derived representations support multiple downstream clinical applications, including risk stratification, improved prognostic performance across cancer types, biomarker characterization and response stratification.

### Spatial reconstruction of biological programs

We first evaluated the ability of SPARC-Map to reconstruct the spatial activity of gene expression programs from H&E images by comparing predicted program scores to the ground truth derived from matched spatial transcriptomics data. This evaluation was performed on 214,933 tissue patches from 71 independent slides in the HEST-Bench [15] dataset. SPARC-Map predictions showed strong concordance with spatial transcriptomics–derived program activity (mean Pearson *r* = 0.74, range 0.49–0.94; mean AUC = 0.90 across programs; range 0.72–0.97), indicating accurate recovery of both continuous variation in program activity (Pearson *r*) and the spatial localization of relatively active versus inactive regions (AUC, based on z-scored program activity).

To contextualize this performance, we compared SPARC-Map to ST-Path [16], an existing method that predicts the spatial expression of individual genes from H&E images. To enable direct comparison at the program level, we aggregated ST-Path gene-level predictions into program scores using the same ssGSEA procedure applied to the ground truth data.

SPARC-Map achieved higher reconstruction fidelity across all 40 biological programs (*P* = 1.8 × 10^−12^, Wilcoxon signed-rank test; Fig. 2(a)). At the individual slide level, SPARC-Map achieved higher mean Pearson correlations in 64 of 71 slides (*P* = 5.2 × 10^−11^; Fig. 2(b)). These results are consistent with the expectation that predicting coordinated programs, rather than individual genes subject to transcript dropout and stochastic noise, yields more stable and biologically coherent spatial reconstructions.

**Figure 2:**
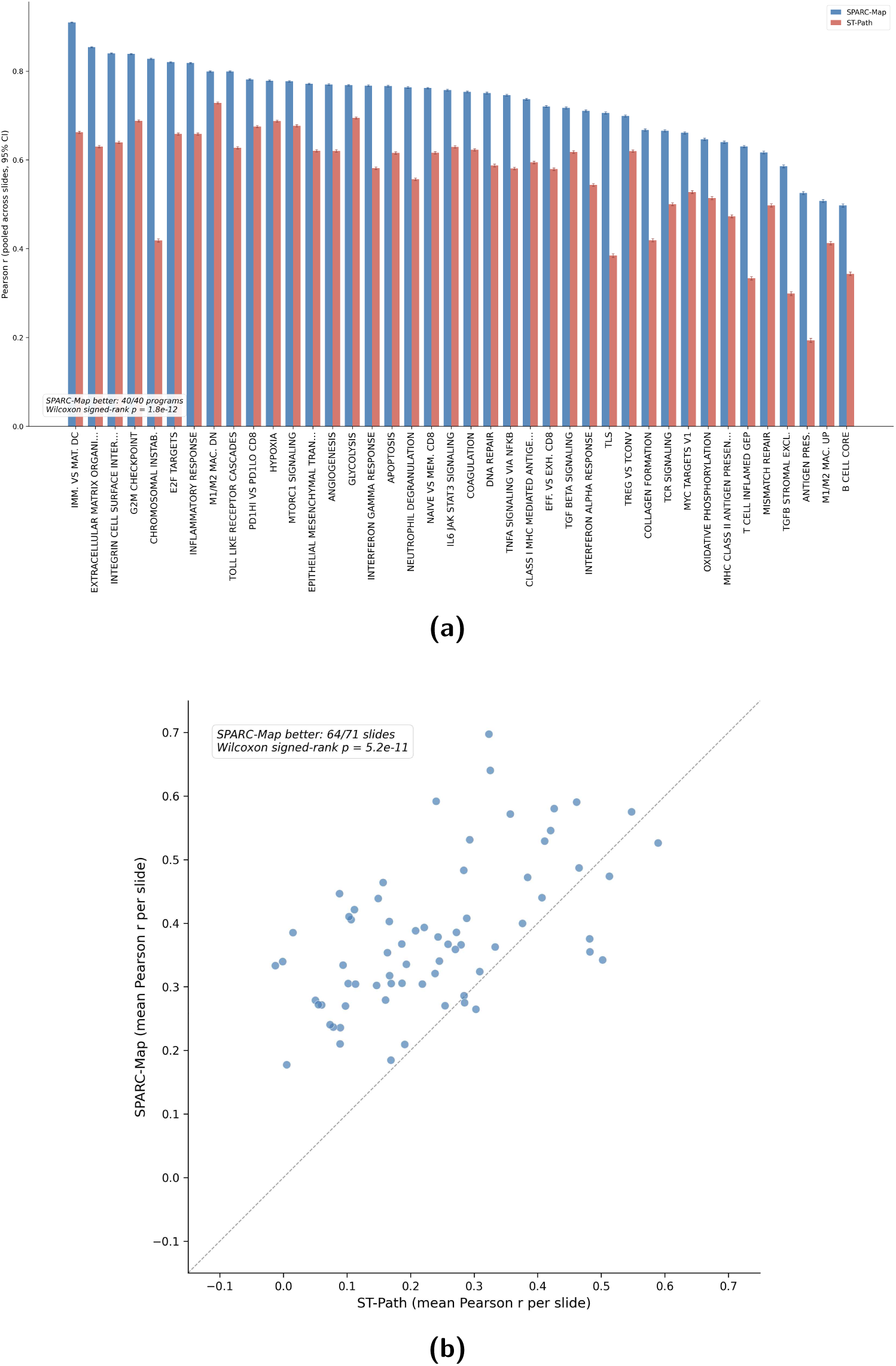
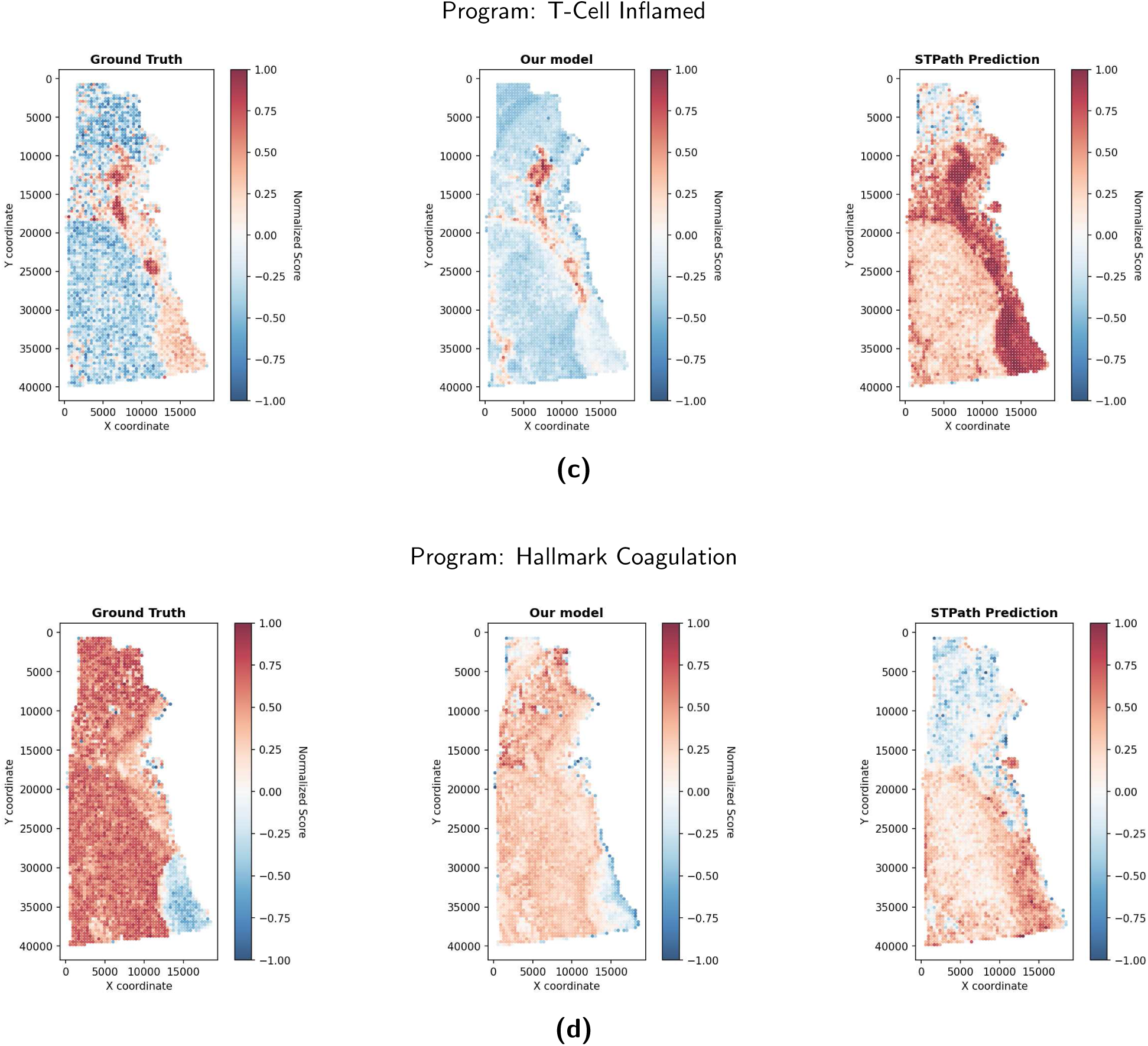
Quantitative benchmarking of SPARC-Map on HEST-Bench. **(a)** Pearson correlation for 40 curated programs pooled across 214,933 tissue patches. SPARC-Map (blue) demonstrates higher concordance than the ST-Path-ssGSEA baseline (red) across evaluated functional domains (*P* = 1.8 × 10^−12^, Wilcoxon signed-rank test). **(b)** Per-slide mean Pearson correlation across 71 independent validation samples. SPARC-Map achieved higher concordance in 64 out of 71 slides (*P* = 5.2 × 10^−11^). (c–d) Representative topographic maps for the T cell-inflamed and coagulation GEPs. Panels show ground-truth spatial transcriptomics (left), SPARC-Map prediction (center), and ST-Path-ssGSEA baseline (right). SPARC-Map preserves architectural boundaries and localized gradients.

Visual inspection of reconstructed maps supported these quantitative findings (Fig. 2(c), 2(d)).

For programs such as T cell-inflamed signaling and the coagulation cascade, SPARC-Map produced sharper, regionally confined patterns that matched ground-truth spatial transcriptomics, whereas gene-level reconstruction produced more diffuse maps, especially in regions affected by transcript sparsity.

### Predicted program maps reveal tumor microenvironment architecture

We hypothesized that spatial co-localization patterns between predicted programs could provide a quantitative, interpretable view of tumor microenvironment (TME) organization.

#### Program-level spatial interactions

We first quantified spatial relationships at the level of individual programs in the TCGA cohort using cross-correlation analysis across tissue space (Fig. 3(a)). This revealed structured and biologically interpretable interactions: TGF-*β* stromal exclusion signatures were strongly anti-correlated with T cell receptor signaling in neighboring patches (*S* = −0.41), consistent with the TGF-*β*-mediated immune exclusion phenotype [17], from H&E alone, while oxidative phosphorylation and glycolysis showed the strongest co-localization (*S* = +0.56), and proliferating tumor regions neighbored exhausted CD8+ T cell zones (*S* = +0.52). All predefined spatial relationships were significant against a permutation null model preserving spatial geometry (*P <* 0.001; Online Methods).

**Figure 3:**
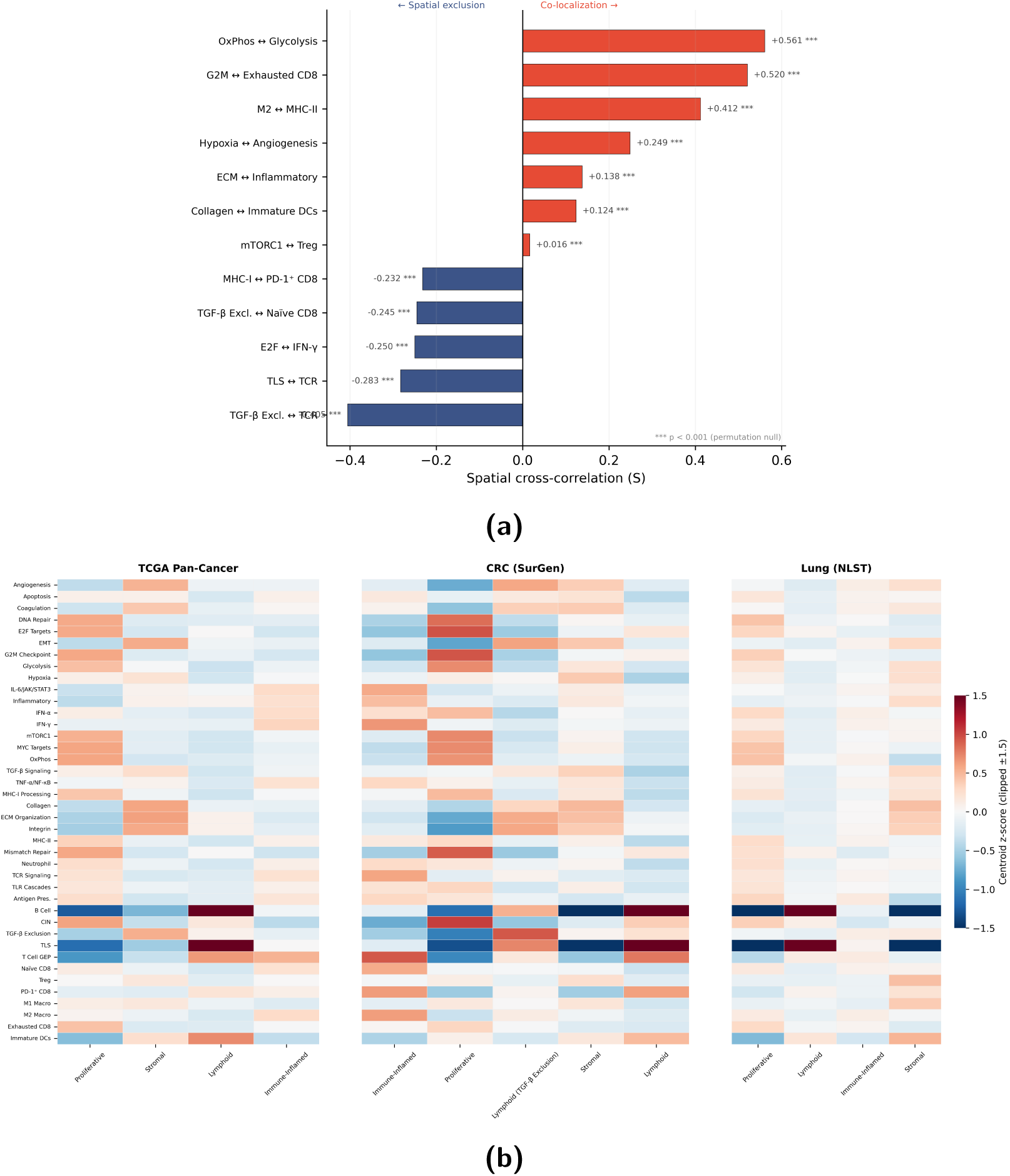
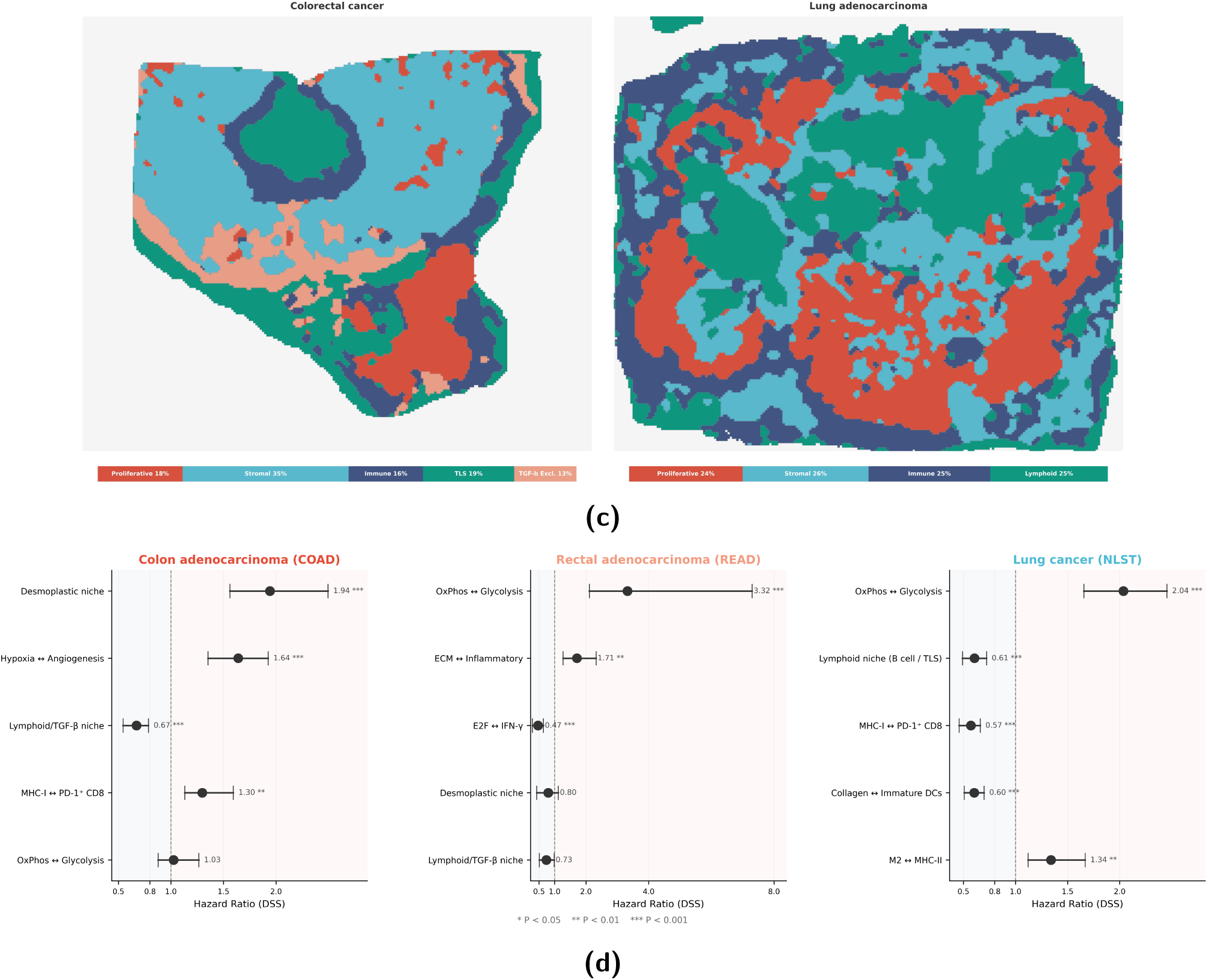
Predicted program maps recover spatial tumor microenvironment ar-chitecture. **(a)** Spatial cross-correlation (*S*) values for predefined biologically motivated program pairs across TCGA. Positive values (red) indicate spatial co-localization; negative values (blue) indicate spatial exclusion. Oxidative phosphorylation and glycolysis show the strongest co-localization (*S* = +0.56), while TGF-*β* stromal exclusion and T cell recep-tor signaling show the strongest anti-correlation (*S* = 0.41). Pairs are sorted by effect size. All pairs are significant against a permutation null model preserving spatial geometry (*P <* 0.001; 200 permutations, 950 slides). **(b)** Niche centroid profiles from unsupervised clustering of SPARC-Map predictions. Left: four pan-cancer niches identified across 8,796 TCGA slides. Center: five disease-specific niches in colorectal cancer (SurGen), revealing a TGF-*β* exclusion niche not resolved in pan-cancer analysis. Right: four niches in lung can-cer (NLST), including an immunoregulatory niche compositionally distinct from the CRC desmoplastic niche. Color encodes centroid value for each program (red, positive enrichment; blue, negative); programs are grouped by functional category along the y-axis. Predicted program maps recover spatial tumor microenvironment ar-chitecture (continued). **(c)** Representative whole-slide niche maps for a colorectal cancer case (SurGen, left) and a lung cancer case (NLST, right). Each patch is colored by its as-signed niche. Insets show the corresponding H&E morphology. The CRC case is consistent with spatial separation between desmoplastic-associated regions (green) and proliferative re-gions (red), while the lung case shows intermixed immunoregulatory and lymphoid regions. Scale bars: 1 mm. **(d)** Cancer-specific prognostic associations of spatial features in external cohorts. Forest plots show hazard ratios (disease-specific survival) for the top spatial fea-tures in SurGen colon cancer (COAD, left), SurGen rectal cancer (READ, center), and NLST lung cancer (right). Each cancer type is driven by a distinct spatial phenotype: desmoplas-tic stroma and hypoxia–angiogenesis coupling in colon cancer, metabolic coupling (OxPhos Glycolysis, HR = 3.32) in rectal cancer, and both metabolic coupling (HR = 2.04, ad-verse) and an immunoregulatory niche (HR = 0.46, protective) in lung cancer. Error bars denote 95% confidence intervals; dashed line indicates HR = 1. \**P <* 0.05, \*\**P <* 0.01, \*\*\**P <* 0.001. Summary results are provided in Supplementary Tables 8 and 9.

#### TME niche organization

To facilitate interpretation of these patterns, we next organized the full set of 40 gene expression programs into a smaller number of program classes using unsupervised k-means clustering across TCGA. The resulting clusters corresponded to four well-recognized niches in the tumor microenvironment—proliferative, stromal, lymphoid, and immune-inflamed compartments (Fig. 3(b))[18–20]—and were highly stable across independent subsamples (centroid cosine similarity *>* 0.999; Supplementary Table 12). Using this coarser representation, we characterized how niche classes are spatially organized across neighboring tissue regions. Certain classes were consistently found in proximity, whereas others were spatially segregated — patterns that are non-trivial given that niche identity was assigned independently at each patch. Together, these neighborhood-level relationships define a ”grammar” of TME organization, in which specific combinations of biological processes preferentially co-occur in space, while others tend to occupy distinct tissue compartments.

#### Disease-specific spatial phenotypes

We then asked whether this spatial organization generalizes across diseases and whether disease-specific spatial phenotypes are associated with clinical outcome. In external cohorts, we summarized program organization into cohort-specific spatial niches using the same unsupervised procedure while allowing the number and composition of clusters to vary. Each cancer type showed a distinct spatial phenotype (Fig. 3(c), 3(d)). In colon cancer, a desmoplastic niche enriched for collagen, extracellular matrix, and epithelial–mesenchymal transition programs was the dominant prognostic feature (HR = 1.94, *P <* 0.001), consistent with CMS4 mesenchymal biology [21]. Rectal cancer was instead driven by spatial co-localization of oxidative phosphorylation and glycolysis (HR = 3.32, *P <* 0.001). In lung cancer, spatial co-localization of oxidative phosphorylation and glycolysis was adverse (HR = 2.04, *P <* 0.001), paralleling the rectal cancer finding (HR = 3.32) and suggesting that spatial metabolic coupling is a shared adverse feature of aggressive tumors across multiple tissue types. In contrast, a lymphoid niche enriched for tertiary lymphoid structure and B cell signatures was protective (HR = 0.61, *P <* 0.001), consistent with published literature linking tertiary lymphoid structures to improved NSCLC outcomes [22, 23]. Both effects were directionally consistent when LUAD and LUSC were analyzed separately (Supplementary Table 10). Representative niche morphology for each cohort is shown in Supplementary Figs. 1 and 2.

#### Context-dependent lymphoid organization

Lymphoid spatial features also showed cancer-specific directionality. TLS TCR co- localization captures the degree to which T cell receptor signaling activity is spatially concentrated within tertiary lymphoid structures rather than dispersed across the tumor. TLS TCR co- localization was adverse in KIRC (HR = 1.60) but protective in HNSC (HR = 0.83), consistent with the documented tissue-specific finding that immune infiltration and lymphoid organization carry inverted prognostic meaning in kidney cancer relative to most solid tumors [24, 25]. Compact spatial survival summaries are provided in Supplementary Tables 7, 8, and 9, and niche multivariable C-indices are summarized in Supplementary Table 11.

Together, these analyses give us a clearer insight into the grammar captured by the model, revealing TME architecture from pairwise program interactions to higher-order niches and linking these organizational patterns to clinical outcomes.

### Pan-cancer prognostic value of program-level features

Next, we evaluated whether spatial patterns in predicted biological program maps provide additional prognostic value beyond standard morphological patch embeddings across cancers in the TCGA pan-cancer cohort. The model achieved a mean C-index of 0.704 across cancer types (range: 0.558–0.864), indicating strong overall prognostic performance.

To assess whether spatial patterns in predicted biological programs provide information beyond histomorphology, we compared the full model to an ablated image-only version using 5-fold cross-validation across 18 evaluable disease groups in the TCGA pan-cancer cohort (COAD and READ merged as colorectal cancer, CRC, for reporting). Cross-validation was used to obtain stable estimates within each cancer type, where sample sizes and event counts are limited despite the overall scale of TCGA.

#### Pan-cancer Discrimination

SPARC-Risk achieved a higher concordance index (C-index, a measure of how well the model ranks patients by risk) in 17 of 18 disease groups (Wilcoxon signed-rank *P* = 1.0 × 10^−4^; Fig. 4(a)) compared to the image-only model. Bootstrap 95% confidence intervals for the difference in C-index (SPARC-Risk minus image-only) showed the largest gains in cervical cancer (CESC: +0.058, 95% CI [−0.004, +0.198]), breast cancer (BRCA: +0.029, [−0.007, +0.058]), clear cell renal cancer (KIRC: +0.029, [−0.008, +0.071]), and bladder cancer (BLCA: +0.022, [−0.007, +0.052]), with only esophageal cancer (ESCA) showing a small decrement (0.009, [0.091, +0.033]). ESCA is histologically heterogeneous, encompassing squamous cell carcinoma and adenocarcinoma subtypes with distinct risk factors, cellular origins, and anatomical distributions [26, 27]. Both histologies carry strong morphological determinants of prognosis, including depth of invasion, tumor differentiation, and lymphovascular invasion, that are independently prognostic and already richly encoded in histopathological images [28–30]. We speculate that these strong morphological signals leave little room for program-level features to improve on.

**Figure 4:**
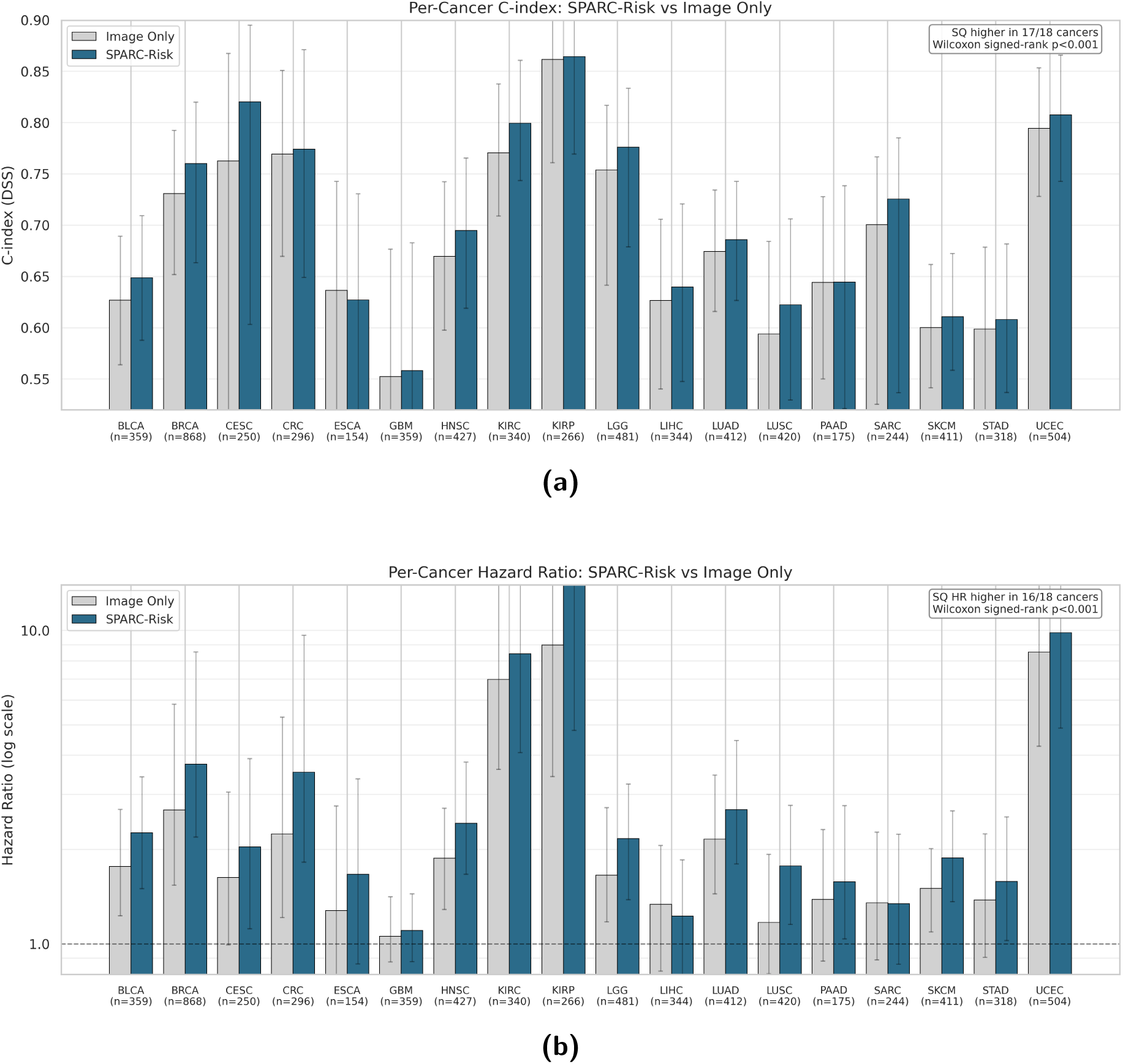
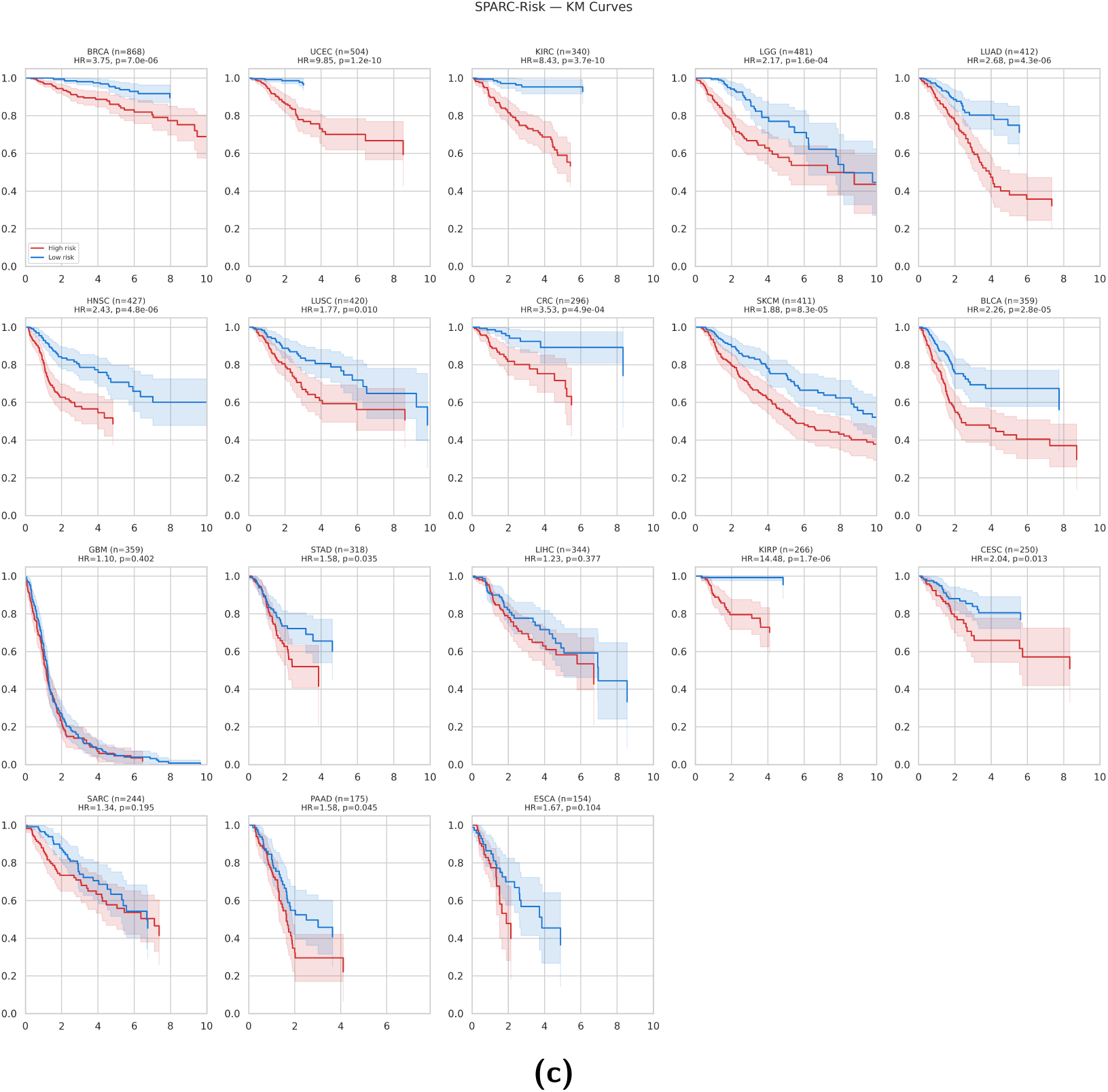
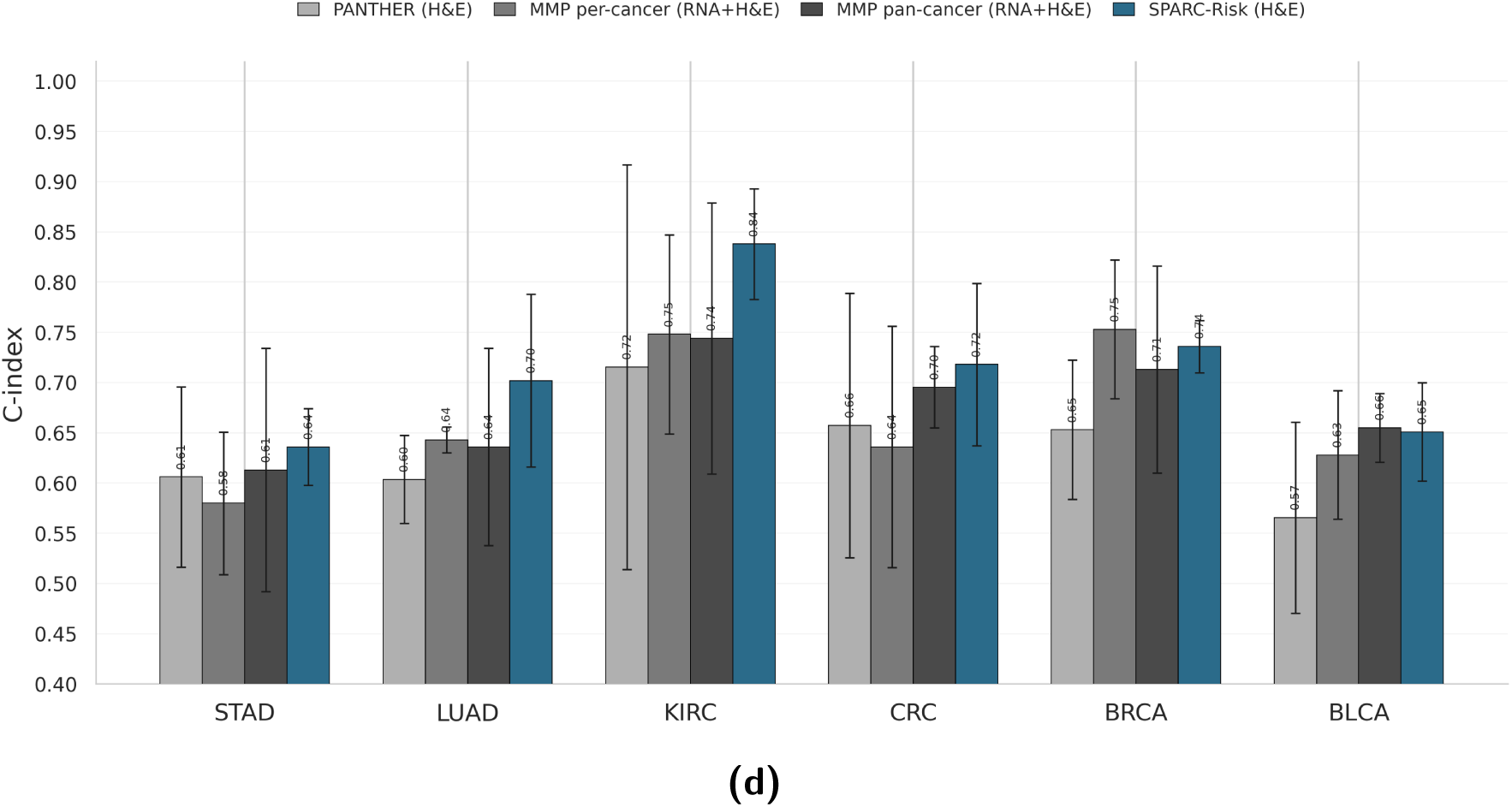
Pan-cancer prognostic utility and benchmarking. **(a)** Bootstrap-estimated C-index differences (ΔC-index; SPARC-Risk minus Image Only) across 18 TCGA disease groups (COAD and READ merged as CRC). Discrimination improved in 17/18 groups (Wilcoxon signed-rank *P* = 1.0 10^−4^); horizontal intervals denote 95% confidence intervals. **(b)** Pan-cancer Hazard Ratio (HR) comparison for high- vs. low-risk groups across the same 18 groups. Using pooled out-of-fold median-split risk stratification per cancer, SPARC-Risk yielded stronger risk separation in 16/18 groups (Wilcoxon signed-rank *P <* 1.0 × 10^−4^). **(c)** Rep-resentative Kaplan–Meier risk stratification from the same pooled out-of-fold, median-split analysis used for panel (b), which showed stronger HR separation for SPARC-Risk in 16/18 disease groups (Wilcoxon signed-rank *P <* 1.0 × 10^−4^). **(d)** Bench-marking against MMP results on standardized test splits in the same six cohorts reported by Song et al. [11] (STAD, LUAD, KIRC, CRC, BRCA, BLCA). Values are mean s.d. across folds for each cancer. Operating unimodally at inference, SPARC-Risk achieved the highest macro C-index (0.71), compared with 0.68 for MMP pan-cancer and 0.63 for PANTHER.

Taken together, these distributed gains across cancer types indicate that program-level features capture prognostic signals not fully represented by morphology alone.

#### Risk stratification

When patients were divided into high- and low-risk groups using median predicted risk scores, SPARC-Risk yielded larger hazard ratios (a measure of the relative risk between groups) in 16 of 18 disease groups (Wilcoxon signed-rank *P <* 1.0×10^−4^; Fig. 4(c), 4(b)) compared to the image-only model. The largest absolute gains in hazard ratio were observed in papillary renal cancer (KIRP: +5.50; 8.98 to 14.48), KIRC (+1.44; 6.99 to 8.43), endometrial cancer (UCEC: +1.31; 8.53 to 9.85), CRC (+1.29; 2.24 to 3.53), and BRCA (+1.07; 2.68 to 3.75). LIHC and SARC showed small decrements (0.11 and 0.01, respectively). Exact per-cancer values are provided in Supplementary Table 3.

We further examined how the contribution of program-level features varies across cancer types. Using SPARC-Risk’s learned per-cancer weights that quantify the relative contribution of predicted biological programs and tissue morphology, we found that the contribution of program-level information was not uniform: cancers with larger performance gains over the image-only model showed greater reliance on program-level features (Supplementary Table 2, Supplementary Fig. 3).

#### Spatial modeling

To assess the contribution of spatial context, we compared our full model, which explicitly captures interactions between each tissue patch and its 64 nearest neighbor patches (k = 64), to a non-spatial variant (*k* = 1) that considers each patch independently. The spatial model outperformed the non-spatial variant in 14 of 18 cancers (*P <* 0.01), indicating that incorporating local tissue context improves prognostic performance.

#### Comparison with multi-omic integration

We compared SPARC-Risk against the MultiModal Panther (MMP) framework, which combines histology with paired bulk RNA sequencing, and its unimodal image-only variant PANTHER [11]. For direct comparability, we restricted this benchmark to the same six cancer cohorts and patient splits reported in the MMP study (STAD, LUAD, KIRC, CRC, BRCA, BLCA) and additionally trained a pan-cancer MMP variant. Across these six cancers (Fig. 4(d)), SPARC-Risk achieved the highest macro-averaged C-index (0.71), exceeding MMP pan-cancer (0.68) and PANTHER (0.63). At the individual cancer level (mean ± standard deviation), SPARC-Risk was the top-performing model in five of six cohorts: STAD (0.64±0.04), LUAD (0.70±0.09), KIRC (0.84±0.06), CRC (0.72±0.08), and BLCA (0.65±0.05), with BRCA near the top and slightly below per-cancer MMP (0.74±0.03 vs. 0.75±0.07). Notably, these results were achieved using only H&E input, whereas MMP requires matched RNA sequencing. This suggests that program-level spatial features extracted from morphology can match or exceed multi-omic baselines in several settings despite relying on H&E alone.

#### Comparison with clinical features

Prior literature has demonstrated that clinical features such as age, sex, and tumor grade have strong predictive ability for patient outcomes [31]. We therefore assessed whether SPARC-Risk provides additional prognostic value beyond these standard variables by fitting multivariable Cox proportional hazards models within each cancer type (Table 1). Across 17 of 18 evaluable cancers, models combining SPARC-Risk with clinical covariates improved discrimination compared to clinical variables alone, indicating that the program adds incremental value. SPARC-Risk remained an independent predictor of survival in 16 of 18 evaluable cancers. The two exceptions were glioblastoma (GBM), where prognosis is dominated by molecular subtypes such as IDH mutation status that are not explicitly captured by our 40-program dictionary [32], and pancreatic cancer (PAAD), which showed a non-significant trend. We next evaluated whether SPARC-Risk adds value beyond established molecular biomarkers with known prognostic strength, such as receptor status in breast cancer (e.g., HER2 or ER). In molecularly annotated subsets, adding SPARC-Risk to models that already included clinical and molecular covariates improved C-index in five of six cancers (mean improvement: +0.109; Supplementary Fig. 4). Per-covariate comparisons are provided in Supplementary Table 4.

**Table 1:** Multivariable clinical adjustment in TCGA.

| Cancer | Cohort Size (Events) | Clinical C-index | SPARC-Risk C-index | Combined C-index | Adjusted HR (95% CI) | P-value |
| --- | --- | --- | --- | --- | --- | --- |
| BLCA | 356 (113) | 0.582 | <u>0.654</u> | **0.675** | 1.64 (1.35–1.98) | < 0.001 |
| BRCA | 868 (60) | 0.584 | <u>0.756</u> | **0.764** | 1.43 (1.23–1.66) | < 0.001 |
| CESC | 246 (48) | 0.583 | <u>0.597</u> | **0.618** | 1.35 (1.05–1.73) | 0.019 |
| COAD | 227 (32) | 0.599 | <u>0.741</u> | **0.757** | 1.80 (1.36–2.39) | < 0.001 |
| ESCA | 154 (46) | 0.614 | <u>0.620</u> | **0.648** | 1.39 (1.01–1.91) | 0.044 |
| GBM | 359 (298) | **0.620** | <u>0.513</u> | **0.620** | 1.01 (0.89–1.14) | 0.905 |
| HNSC | 423 (120) | 0.525 | <u>0.653</u> | **0.657** | 1.61 (1.35–1.92) | < 0.001 |
| KIRC | 337 (58) | 0.757 | <u>0.807</u> | **0.845** | 1.88 (1.48–2.39) | < 0.001 |
| KIRP | 264 (27) | 0.580 | <u>0.890</u> | **0.902** | 3.02 (2.32–3.93) | < 0.001 |
| LGG | 480 (103) | <u>0.788</u> | 0.689 | **0.797** | 1.34 (1.12–1.60) | 0.002 |
| LIHC | 339 (75) | 0.493 | **0.581** | <u>0.569</u> | 1.38 (1.11–1.71) | 0.004 |
| LUAD | 402 (96) | <u>0.534</u> | **0.651** | **0.651** | 1.57 (1.33–1.86) | < 0.001 |
| LUSC | 415 (82) | 0.529 | <u>0.617</u> | **0.636** | 1.30 (1.07–1.56) | 0.007 |
| PAAD | 175 (79) | <u>0.602</u> | 0.584 | **0.620** | 1.26 (0.99–1.60) | 0.056 |
| READ | 69 (5) | — | — | — | — | — |
| SARC | 244 (78) | 0.584 | <u>0.608</u> | **0.640** | 1.31 (1.05–1.63) | 0.019 |
| SKCM | 411 (162) | <u>0.588</u> | 0.586 | **0.620** | 1.28 (1.10–1.48) | 0.001 |
| STAD | 315 (84) | 0.554 | <u>0.589</u> | **0.599** | 1.39 (1.11–1.73) | 0.004 |
| UCEC | 497 (54) | 0.728 | <u>0.787</u> | **0.807** | 1.54 (1.28–1.85) | < 0.001 |

Best C-index per cancer is shown in bold and the second best is underlined. Adjusted HR corresponds to the SPARC-Risk term (risk<sub>z</sub>) in the combined model; cancers with fewer than 15 events were not evaluated.

### External validation across independent cohorts

We evaluated generalizability across three external cohorts from distinct institutions and organ systems: the National Lung Screening Trial (NLST; 422 patients, 1,170 slides)[33], the SurGen colorectal cohort (425 patients, 425 slides)[34], evaluated as separate colon and rectal cancer subsets, and an institutional gastrointestinal cohort from Massachusetts General Hospital (MGH; 94 patients, 127 slides). We report both zero-shot performance (TCGA-trained models applied without adaptation) and a fine-tuned setting, in which the learned representations were held fixed and a ridge-regularized Cox model was fit on the SPARC-Risk embeddings within each cohort.

#### Zero-shot transfer

Without any cohort-specific adaptation, SPARC-Risk outperformed the image-only baseline across all 8 cohort–endpoint combinations (Fig. 5(a)), with a mean C-index improvement of +0.024. In NLST (*n* = 422), gains were consistent for both DSS (0.64 [0.58–0.69] vs. 0.63 [0.58–0.68]) and OS (0.62 [0.57–0.67] vs. 0.61 [0.56–0.66]). In SurGen-COAD (*n* = 260), SPARC-Risk improved both DSS (0.73 [0.67–0.79] vs. 0.71 [0.65– 0.78]) and OS (0.68 [0.62–0.72] vs. 0.65 [0.59–0.70]). The largest gains were observed in SurGen-READ (*n* = 165; DSS: 0.63 [0.53–0.72] vs. 0.57 [0.50–0.67]; OS: 0.57 [0.51–0.65] vs. 0.54 [0.50–0.62]). In MGH GI (*n* = 94), SPARC-Risk improved DSS (0.73 [0.66–0.81] vs. 0.72 [0.65–0.80]) and OS (0.74 [0.66–0.81] vs. 0.72 [0.64–0.79]).

**Figure 5:**
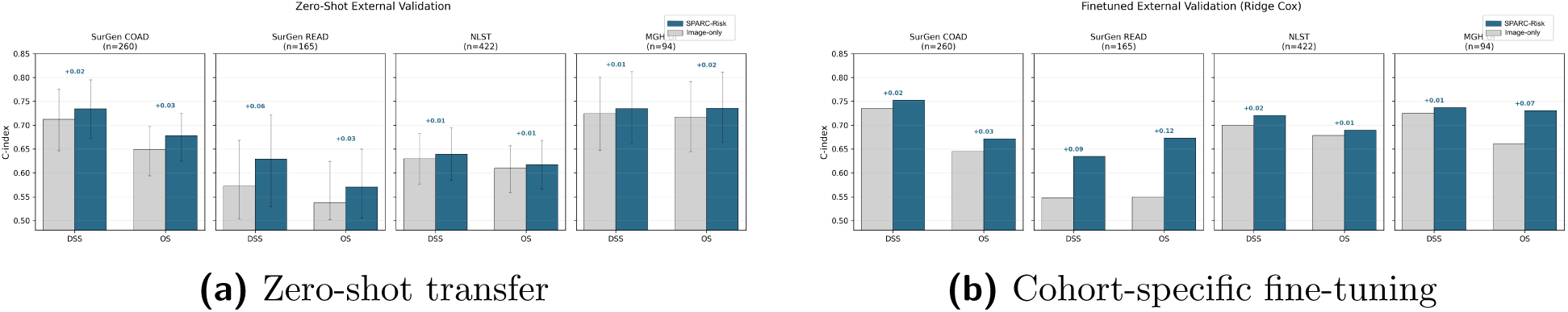
External validation across independent cohorts in zero-shot and fine-tuned settings. **(a)** Zero-shot C-index comparisons (SPARC-Risk vs. image-only) for DSS and OS endpoints in NLST, SurGen-COAD, SurGen-READ, and MGH GI; brackets indicate 95% confidence intervals. **(b)** Corresponding C-index comparisons after cohort-specific fine-tuning on the same cohort-endpoint pairs.

#### Fine-tuned performance

After cohort-specific fine-tuning, SPARC-Risk again exceeded the image-only baseline for all 8 endpoints (Fig. 5(b)), with a larger mean improvement of +0.046. In NLST, SPARC-Risk improved DSS (0.72 vs. 0.70) and OS (0.69 vs. 0.68). In SurGen-COAD, SPARC-Risk improved DSS (0.75 vs. 0.73) and OS (0.67 vs. 0.64). The largest gains were in SurGen-READ, where SPARC-Risk exceeded the image-only baseline for both DSS (0.63 vs. 0.55; +0.09) and OS (0.67 vs. 0.55; +0.12). In MGH GI, a notable +0.07 OS gain was observed (0.73 vs. 0.66). The consistent improvement from zero-shot to fine-tuned settings indicates that the learned representation is both transferable and readily adaptable with modest cohort-specific data.

### Stratification of therapeutic response

To test whether predicted program maps carry signals relevant to treatment selection, we applied SPARC to two retrospective targeted-therapy cohorts: ovarian cancer treated with bevacizumab plus chemotherapy (bevacizumab is an anti-angiogenic agent; *n* = 73 patients, 252 slides)[35, 36] and breast cancer treated with trastuzumab plus chemotherapy (trastuzumab is an anti-HER2 agent; *n* = 85 patients, 85 slides)[37, 38]. In each cohort, we evaluated supervised stratification (logistic regression on predicted program scores) and unsupervised stratification (clustering without outcome labels), benchmarking against image-only features. Across both cohorts, program-based features outperformed image-only baselines in both settings.

#### Ovarian cancer: bevacizumab response

The cohort is predominantly high-grade serous (74% of patients), with the remainder comprising endometrioid, clear cell, mucinous, and unclassified adenocarcinoma subtypes [35]. Response was defined by CA-125 criteria combined with imaging-based assessment of tumor progression within six months of treatment, serving as a surrogate for clinical benefit rather than a direct survival endpoint. Program-based stratification was significant in both the supervised and unsupervised settings (supervised odds ratio, OR = 8.08, 95% CI [2.71, 24.05], *P* = 1.16×10^−4^; unsupervised OR = 5.77, 95% CI [1.94, 17.21], *P* = 0.0013; Fig. 6(a)). Image-only baselines did not reach significance in either setting (supervised OR = 2.77, 95% CI [1.00, 7.69], *P* = 0.066; unsupervised OR = 2.49, 95% CI [0.92, 6.75], *P* = 0.087). Notably, significance was observed with both supervised modeling and unsupervised clustering, indicating that the response signal was not limited to a single stratification strategy. Feature-ablation results including baseline CA-125 levels are provided in Supplementary Table 6.

**Figure 6:**
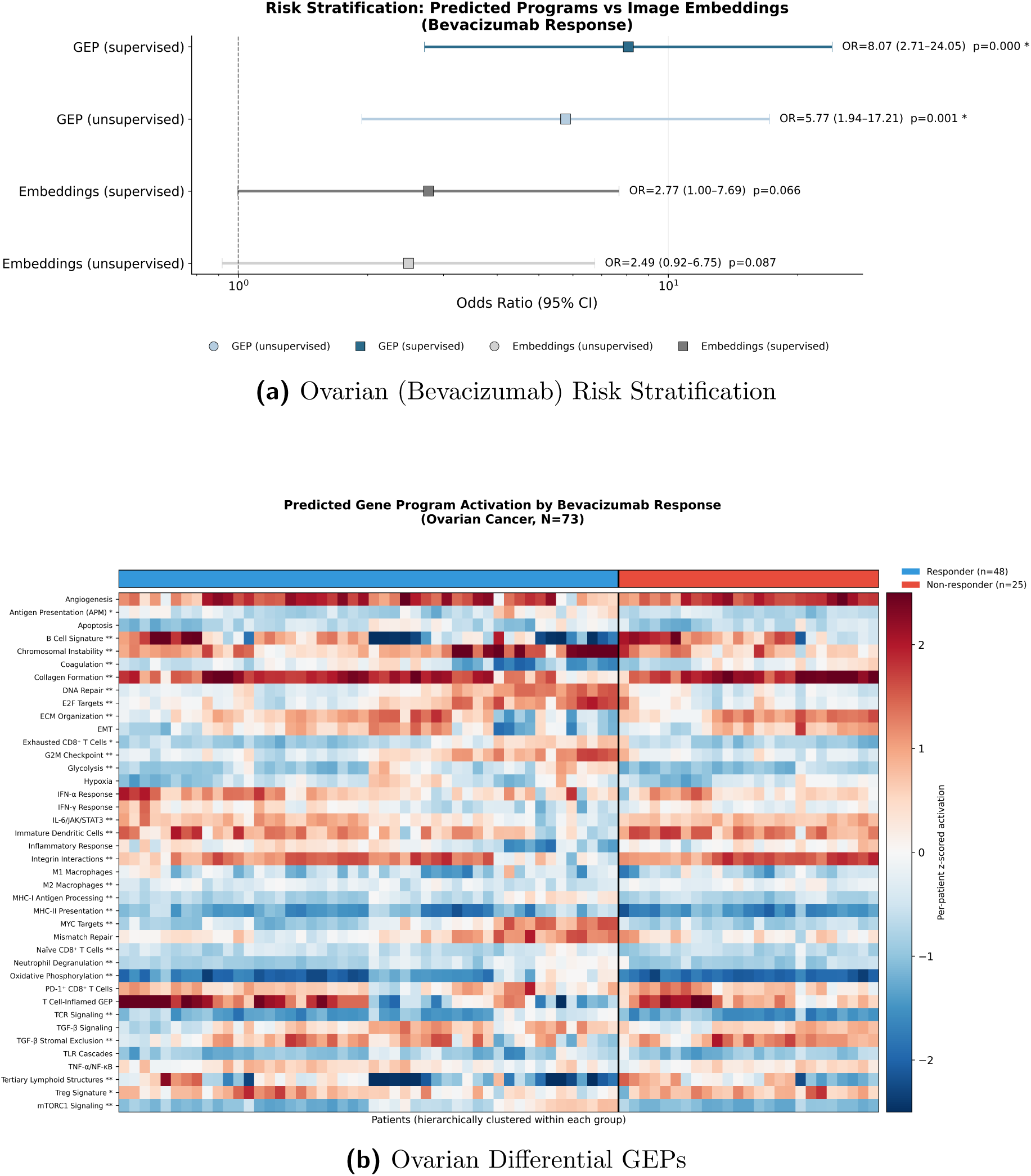
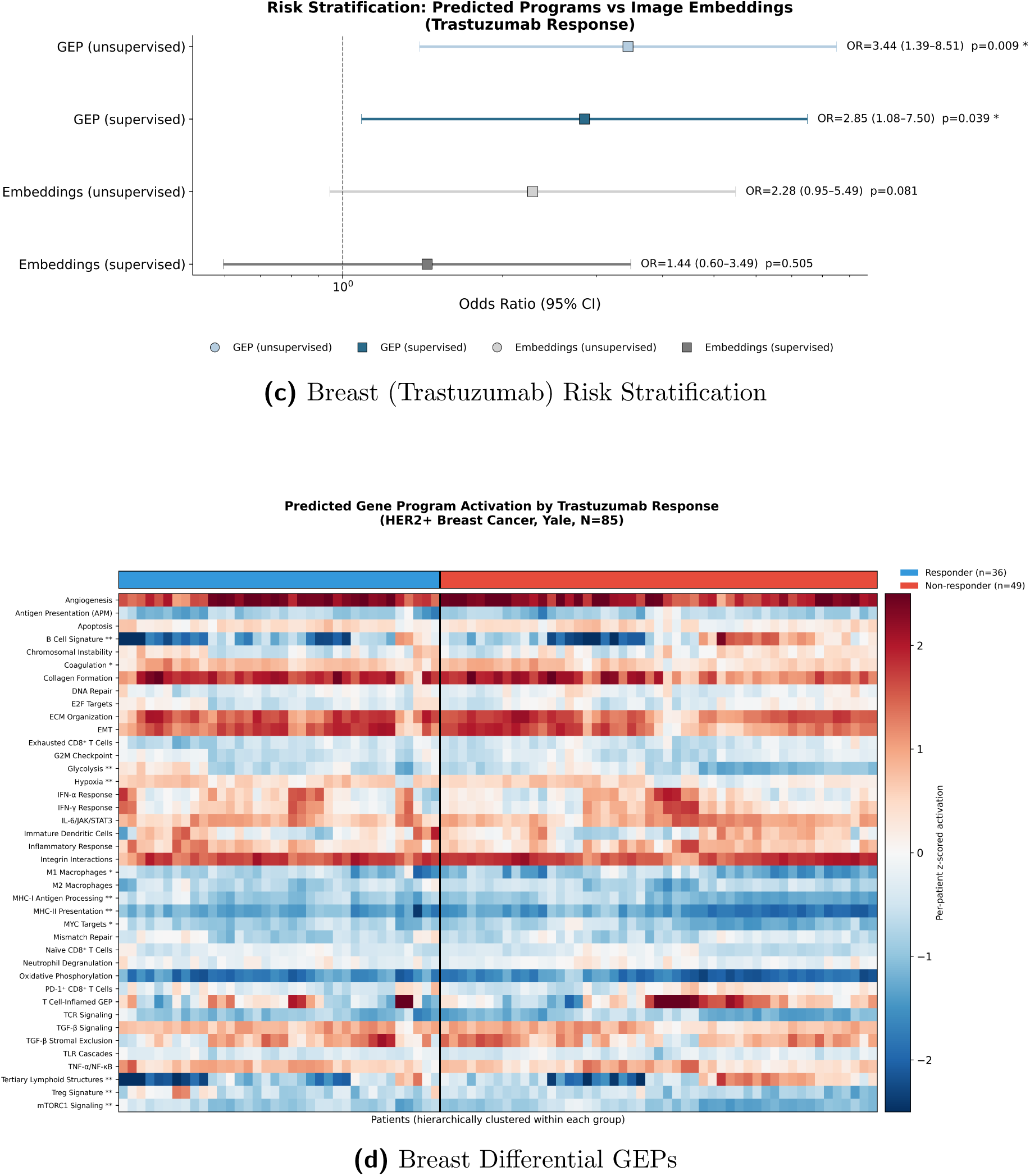
Mechanism-aware prediction of targeted therapy response. **(a)** Risk stratification for bevacizumab response in the ovarian cancer cohort (*n* = 73). GEP features stratify response in both supervised (OR = 8.08, *P* = 1.16 × 10^−4^) and unsupervised (OR = 5.77, *P* = 0.0013) settings, whereas image-only baselines are weaker and non-significant (supervised OR = 2.77, *P* = 0.066; unsupervised OR = 2.49, *P* = 0.087). **(b)** Topographic differential program activation in the ovarian cohort. The SPARC-Map predictions identify 26 nominally significant programs, with 23 retained after both BH-FDR and permutation-FDR correction. Differential signal spans proliferative/metabolic and adaptive immune programs in responders, and stromal/immunoregulatory programs in non-responders. **(c)** Risk stratification for trastuzumab response in the data-restricted breast cancer cohort (*n* = 85). GEP features stratify response in both supervised (OR = 2.85, *P* = 0.039) and unsupervised (OR = 3.44, *P* = 0.0085) settings, whereas image-only baselines are weaker and non-significant (supervised OR = 1.44, *P* = 0.505; unsupervised OR = 2.28, *P* = 0.081). **(d)** Differential program activation in the breast cancer cohort from per-patient z-scored GEPs. Eight of 40 programs are significant after FDR correction, with responder enrichment for antigen-presentation/metabolic modules and non-responder enrichment for TLS/B-cell signatures.

#### Breast cancer: trastuzumab response

In the smaller trastuzumab cohort (85 slides vs 252 in the ovarian cohort), both supervised and unsupervised program-based analyses reached significance (supervised OR = 2.85, 95% CI [1.08, 7.50], *P* = 0.039; unsupervised OR = 3.44, 95% CI [1.39, 8.51], *P* = 0.0085; Fig. 6(c)). Image-only baselines did not reach significance (supervised OR = 1.44, 95% CI [0.60, 3.49], *P* = 0.505; unsupervised OR = 2.28, 95% CI [0.95, 5.49], *P* = 0.081). Similarly to the ovarian setting, both analyses remained significant despite the smaller cohort, suggesting that the program-derived signal is relatively robust in this setting.

## Biological interpretation of therapy response

### Ovarian cancer

Differential analysis revealed broad differences between response groups: 26 of 40 programs were nominally significant, and 23 remained significant after both BH-FDR (Benjamini–Hochberg false discovery rate [39]) and permutation FDR correction (Fig. 6(b)). The resulting profiles aligned with independently reported molecular subtypes. Responders showed higher activity in a coherent proliferative and metabolic program—oxidative phosphorylation, glycolysis, mTORC1 signaling, E2F targets, G2/M checkpoint, MYC targets, DNA repair, and chromosomal instability (FDR *<* 0.05)—consistent with ICON7 analyses reporting greater bevacizumab benefit in tumors with proliferative molecular subtypes [12]. Non-responders instead showed higher stromal and immunoregulatory activity, including M2 macrophage and immature dendritic cell signatures, IL-6/JAK/STAT3 and TGF-*β* Stromal Exclusion, extracellular matrix organization, collagen formation, and integrin interactions (FDR *<* 0.05), consistent with a fibrotic, immune-excluded microenvironment that may be less dependent on VEGF-driven angiogenesis. This pattern is also consistent with MITO16A/MaNGO-OV2, which linked stromal and immunoregulatory features to poorer bevacizumab outcomes [40]. Notably, adaptive immune programs (T cell receptor signaling, MHC class I and class II antigen presentation, and näıve CD8^+^ T cell signatures) were also higher in responders, suggesting that the adverse biology in non-responders is concentrated in immunosuppressive myeloid and stromal pathways rather than reduced adaptive immunity per se.

### Breast cancer

After per-patient z-scoring, differential analysis identified 8 of 40 programs with significant responder/non-responder differences (FDR *<* 0.05; Fig. 6(d)), with responders relatively enriched for antigen-presentation and metabolic programs and non-responders for adaptive immune structural programs, including tertiary lymphoid structures and B cell signatures. More importantly, raw program scores (without per-patient z-scoring) showed globally higher activity in responders across nearly all programs (38 of 40 higher; Mann–Whitney U test [41] *P* = 0.009; Supplementary Fig. 5), indicating that the dominant distinction in this cohort is a broad pan-active responder state. This pattern is consistent with HER2-enriched biology and prior studies linking broad immune and metabolic activation to trastuzumab benefit [42–44]. In supporting analyses (Supplementary Table 5), program-based features outperformed receptor-status clinical variables and image-only features (AUC 0.588 vs. 0.451 and 0.561), whereas clinical variables alone were not predictive (OR = 1.15, *P* = 0.8141). Combining clinical variables with program features performed best (AUC 0.623, OR = 3.38, 95% CI [1.24, 9.17], *P* = 0.0203). Program-based clustering also stratified responders within the ER-positive subgroup (71% vs. 35%; P=0.030), where ER status itself did not distinguish responders from non-responders, suggesting that program-level features capture response-relevant biology beyond what receptor status encodes.

## Discussion

In this study, we introduced SPARC, a framework that bridges the gap between clinical outcome prediction from H&E images and the underlying molecular mechanisms that drive disease progression. SPARC produces interpretable maps showing where specific processes are predicted to be active across the tissue, providing a biological readout that can be interrogated and compared against known molecular phenotypes. This program-level representation improved prognostic performance over image-only models in 17 of 18 cancer types, demonstrating that the additional interpretability does not come at the cost of predictive accuracy. Beyond prognostic improvement, unsupervised analysis of predicted program maps recovered canonical tumor microenvironment compartments and spatial interaction patterns, including TGF-*β*-mediated immune exclusion architecture, directly from H&E morphology. Disease-specific spatial analyses revealed that each cancer type is characterized by a distinct dominant spatial phenotype—desmoplastic stroma in colon cancer, metabolic coupling in rectal cancer, and a protective lymphoid niche enriched for tertiary lymphoid structures and B cell signatures in lung cancer—demonstrating that SPARC identifies cancer-type-specific spatial vulnerabilities from routine histology.

A growing body of work has sought to link routine histopathology with molecular information, of which three directions are particularly relevant here. One consists of pathology-only methods that infer spatial transcriptomic signals directly from H&E images [16][45]. These studies show that morphology encodes gene expression signal, but gene-level prediction remains modest; for example, STPath reports an average Pearson correlation of 0.266 across the top 200 highly variable genes, with performance varying across datasets and tissue types. A second direction predicts spatial proteomic features from histology, including virtual multiplexed immunofluorescence approaches such as GigaTIME [46], which enable cell-level phenotyping but are constrained by predefined antibody panels. A third direction integrates histology with paired molecular data, such as bulk or spatial RNA sequencing [11][47], improving predictive performance but requiring matched datasets and, for bulk RNA sequencing, sacrificing spatial resolution. Related H&E-only studies [48] have also shown that tumor-level molecular states, such as integrative subtypes or relapse-risk categories, can be predicted directly from histology, but these approaches operate at the level of global endpoints rather than spatially resolved biological representations. In contrast, our framework is also H&E-only but represents tissue in terms of coordinated biological programs and their spatial organization rather than individual molecular features. We find that this approach performs comparably to multimodal frameworks requiring paired RNA sequencing and histology, while capturing prognostic information that can complement—and in some settings partially substitute for—bulk molecular profiling.

Beyond prognosis, the therapy-response analyses illustrate how this approach can connect predictions to clinical decision making. In both the ovarian and breast cancer cohorts, program-based features achieved stronger and statistically significant stratification of responders and non-responders, whereas image-only features did not reach significance. Importantly, the biological profiles identified by SPARC aligned with independently reported molecular subtypes: proliferative programs predicted bevacizumab benefit in ovarian cancer, and broad immune-metabolic activation predicted trastuzumab response in breast cancer. These alignments were not built into the model; they emerged from the predicted program scores, suggesting that SPARC recovers biologically meaningful signals rather than spurious morphological correlations. Furthermore, unsupervised clustering, which requires no outcome labels, achieved the strongest stratification in the breast cancer cohort, demonstrating that the biological structure captured by SPARC can stratify patients even without task-specific training. This property is particularly valuable in low-data regimes such as early-phase clinical trials, where supervised approaches are prone to overfitting.

From a translational perspective, SPARC can serve two complementary roles. In settings where molecular assays are unavailable, delayed, or infeasible, which remains common in routine clinical practice, it can support risk stratification, biomarker discovery, and hypothesis generation from the H&E slides already collected as part of routine care. In settings where molecular data are available, it can provide spatial context for pathway activity that bulk measurements cannot capture. By requiring only a standard pathology slide at inference, this framework lowers the barriers to mechanism-informed precision oncology. A natural next step will be to move beyond program-level summaries toward more specific therapeutic hypotheses, ultimately linking histology-derived biology to actionable and potentially druggable targets.

This study has several limitations. First, SPARC relies on a curated dictionary of 40 transcriptomic programs that, while spanning major cancer hallmarks, does not represent the full molecular complexity of tumor evolution. Expanding this framework to additional program sets, or to other molecular modalities such as spatial epigenomics, represents an important direction. Second, SPARC currently operates at the patch level, so each prediction reflects a local aggregate tissue state rather than true single-cell resolution. This localized bulk view may dilute signals carried by rare cell populations, cell states, or fine-grained cell–cell interactions that occur within a patch. Improving the resolution of these representations, potentially through cell-aware modeling or higher-resolution spatial molecular supervision, is therefore an important direction for future work. Third, our external validations are retrospective; prospective evaluation in clinical workflows will be needed to establish clinical utility. Fourth, gains were not uniform across cancer types: esophageal cancer was the only disease group with a small decrement versus image-only prediction (ΔC-index = −0.009), and glioblastoma showed no independent association in multivariable adjustment (adjusted hazard ratio = 1.01, *P* = 0.905). Similarly, program-level features provided limited additional value in the SurGen rectal cancer cohort, suggesting that in some cancers with well-defined tissue architecture, pretrained image features may already capture the key morphological patterns associated with disease progression. These boundary cases motivate the development of adaptive or tissue-specific program dictionaries that can be tailored to the biology of individual cancer types.

## Online Methods

### Data and cohort descriptions

#### SPARC-Map evaluation (HEST-Bench)

We evaluated SPARC-Map on HEST-Bench, a curated subset of the HEST-1k dataset separate from our training data comprising 71 human slides and 214,933 image patches [15]. The cohort spans ten cancer types: CCRCC, COAD, HCC, IDC, LUNG, LYMPH IDC, PAAD, PRAD, READ and SKCM. One slide (TENX99) was excluded from all analyses because its unusually large patch count (20,761 patches) exceeded the available memory required for inference with the ST-Path model. No additional patch filtering was applied. Cohort characteristics are summarized in Supplementary Table 1.

#### Training and internal evaluation (TCGA)

We utilized 8,835 whole-slide images (WSIs) from 7,284 patients across 19 TCGA cancer types [14]: BLCA, BRCA, CESC, COAD, ESCA, GBM, HNSC, KIRC, KIRP, LGG, LIHC, LUAD, LUSC, PAAD, READ, SARC, SKCM, STAD, and UCEC. These cohorts were selected based on a minimum threshold of 100 patients and 20 clinical events, with READ included for comparability with prior TCGA pan-cancer studies despite fewer events (15 events across 152 patients). Patient-level 5-fold cross-validation splits were implemented using hybrid assignments that incorporate standard test-set designations, ensuring all slides from a single patient were restricted to the same split to prevent data leakage.

#### Independent external validation cohorts

To assess real-world generalizability, we evaluated the model on: (i) the NLST lung cancer cohort (422 patients, 1,170 slides) [33]; (ii) the SurGen colorectal cohort (425 patients, 425 slides) drawn from 1,020 WSIs, with survival available for 427 and two cases excluded for unclear cancer type [34]; and (iii) an expanded MGH gastrointestinal (GI) cohort (*n* = 94 patients, 127 slides).

#### Therapeutic cohorts

Therapeutic response was modeled in a Yale HER2+ Breast Cancer cohort (*n* = 85 patients) evaluating Pathologic Complete Response (pCR) to neoadjuvant trastuzumab [38], and an Ovarian Cancer cohort (*n* = 73 patients, 252 slides) evaluating bevacizumab response; in both cohorts, the study agent was administered in combination with chemotherapy.

### SPARC-Map: spatial transformer for program activity prediction

#### Biological program scoring

To translate gene-level expression into robust, biologically interpretable targets for model training, we applied single-sample Gene Set Enrichment Analysis (ssGSEA) [49] to matched spatial transcriptomics data. ssGSEA scores the coordinated expression of predefined gene sets within an individual sample, producing a continuous enrichment score for each program at each spatial location. This approach captures the joint activity of tens to hundreds of genes, reducing sensitivity to the dropout and stochastic noise that affect individual transcript measurements. We curated 40 biological programs from the MSigDB Hallmark collection and Reactome pathway signatures [5], spanning processes including proliferation (for example, E2F targets, G2/M checkpoint), immune signaling (for example, interferon response, antigen presentation), metabolism (for example, oxidative phosphorylation, glycolysis), and microenvironmental remodeling (for example, extracellular matrix organization, angiogenesis). Gene identifiers were standardized to Ensembl Stable IDs. ssGSEA was executed with rank-based normalization, a minimum gene set size of 5, and a maximum size of 5,000.

SPARC-Map (Supplementary Fig. 6) predicts program activity scores for each tissue patch by jointly encoding the patch and its *k* = 16 nearest neighbors using a transformer architecture. Image features extracted from each region are combined with relative spatial encodings, allowing information to be integrated across the local neighborhood via multi-head self-attention. The model consists of an input projection layer followed by three transformer encoder blocks (8 attention heads, hidden dimension *H*= 256). Relative spatial coordinates (Δ*x,* Δ*y*) between each patch and its neighbors are encoded using a multilayer perceptron (2 → 64 → *H*) and incorporated as positional encodings, enabling the model to capture spatial relationships between tissue regions. The model is trained on the HEST-1k dataset using a combined loss *L* = *L*_MSE_ +*λL*_Pearson_ (*λ* = 0.5) with AdamW optimization (*lr* = 10^−4^).

### SPARC-Risk: residual gated fusion and survival modeling

SPARC-Risk integrates predicted program maps with morphological features using a residual gated fusion strategy that allows the model to learn how much additional prognostic value the biological programs provide beyond morphology alone, separately for each cancer type.

#### Dual-pathway architecture

The model comprises two parallel processing streams. The *Image Pathway* projects patch-level Hoptimus1 embeddings (*d* = 1536) [13] to a 256-dimensional representation. The *Biological Enrichment Pathway* takes the 40 predicted program scores, expands them into query vectors (*Q*), and uses multi-head cross-attention to integrate program activity with the morphological features of spatially neighboring patches.

#### Controlled fusion via learned scalar gates

The two pathways are combined using a learned per-cancer-type scalar gate *σ*(*θ*_c_) initialized at *σ*(0) = 0.5:

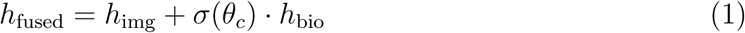

If biological programs provide no additional prognostic value for a given cancer type, the model learns to close the gate (*σ* → 0), effectively defaulting to the image-only baseline. This design ensures that program-level features are incorporated only when they improve prediction.

#### Image-only control baseline

To isolate the contribution of biological programs, we compared SPARC-Risk against a capacity-matched image-only baselines (matched to the depth of SPARC-Risk’s image encoder).

### Spatial analysis of predicted program maps

To assess whether predicted program maps encode biologically interpretable spatial organization, we performed spatial analyses on predicted SPARC-Map outputs across the TCGA pan-cancer cohort and the two external cohorts used for survival validation (SurGen and NLST).

#### Preprocessing

For each slide, predicted program scores were standardized to mean zero and unit variance across all patches (within-slide z-scoring). A per-patch centering step then subtracted the mean across all 40 programs for each patch, removing a shared global activation component that otherwise dominated inter-program correlations (first principal component explained 60–84% of variance across slides; after centering, mean cross-program correlation decreased from 0.77 to approximately 0.02).

#### Spatial graph construction

Patch adjacency was defined using radius neighbors (cKDTree) with an 8-connected radius scaled to the patch step size (radius = patch step × 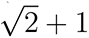), yielding approximately 8 neighbors per patch on average.

#### Spatial cross-correlation

For each slide, a neighbor mean matrix was computed via sparse matrix multiplication, where each entry represents the mean value of a given program across spatial neighbors of a given patch. Spatial cross-correlation *S*(*g*_1_*, g*_2_) was defined as the Spearman correlation between program *g*_1_ at a patch and the mean of program *g*_2_ in its spatial neighborhood. Values were Fisher-z-transformed before averaging across slides. Significance was assessed against a permutation null model (200 permutations across 950 slides, 50 per cancer type) in which patch-program assignments were shuffled while preserving spatial geometry.

#### Spatial niche discovery

Approximately 4.4 million patches were subsampled (capped at 500 per slide) and reduced to 8 principal components (retaining ≥ 90% variance). K-Means clustering was performed for *K* 4, 5, 6, 7, 8 with 50 initializations each; the best *K* was selected by silhouette score. Frozen centroids were then used to assign all patches across all slides by nearest-centroid distance. Patient-level niche composition was computed as the fraction of patches assigned to each niche, with Shannon entropy summarizing niche diversity. For multi-slide patients (NLST), niche counts were summed across slides before computing fractions. Disease-specific clustering (*K* = 5 for SurGen, *K* = 4 for NLST) was performed separately on each external cohort.

#### Spatial prognostic analysis

Univariate Cox proportional hazards models were fit for each spatial feature (pairwise *S* scores, niche proportions, entropy) within each disease group. Abundance-adjusted models additionally included mean program levels for both programs in a pair, testing whether spatial organization predicts survival beyond program abundance. All survival analyses used disease-specific survival as the primary endpoint. Compact TCGA and external-cohort spatial survival summaries are reported in Supplementary Tables 7–9.

### Clinical validation and downstream analyses

#### External survival validation

All splits were patient-level. Model risk scores were generated using 5-fold cross-validation, and reported metrics use fold-aggregated predictions (or pooled-fold analyses where noted). Evaluation followed two regimes: (i) Zero-shot, averaging predictions from the five TCGA fold models; and (ii) Ridge Cox optimization, utilizing *L*_2_-regularized Cox models on slide embeddings. For the latter, *λ* was selected via inner 3-fold cross-validation over 10^−4^*, …,* 100.0, and final performance was reported using out-of-fold predictions from an outer 5-fold stratified cross-validation. Fine-tuning therefore amounts to fitting a Cox linear model on the final SPARC-Risk embeddings, which is equivalent to learning only the risk head while keeping upstream representations frozen. Hyperparameters were fixed a priori and applied uniformly across cohorts.

#### TCGA pan-cancer bootstrap comparison

For the internal TCGA comparison in Fig. 4(a) and Fig. 4(b), we benchmarked SPARC-Risk versus the image-direct baseline across 18 disease groups (COAD and READ merged as CRC). Per-cancer C-indices were computed within each held-out fold from 5-fold cross-validation and averaged across folds for point estimates. For hazard-ratio analysis, patients were stratified into high- and low-risk groups using the median predicted risk score, and HRs (high vs. low) were computed from pooled out-of-fold predictions within each disease group using log-rank estimation. This pooling was used to stabilize HR estimation under limited per-fold event counts while preserving out-of-sample evaluation because each patient prediction was generated by a model that did not observe that patient’s outcome during training. Bootstrap confidence intervals (95%; 1,000 paired patient-level nonparametric resamples) were computed by resampling the same patients simultaneously for both models. Overall cross-cancer significance was assessed using Wilcoxon signed-rank tests on the 18 per-cancer differences (ΔC-index or ΔHR).

Therapeutic response modeling. To evaluate the robustness of program-based features for treatment-response prediction, we applied both supervised and unsupervised strategies to both therapeutic cohorts.

- **Supervised linear probing**: For each cohort, a logistic regression model was trained on patient-level SPARC-Map representations (mean of patch-level program scores) to assess discriminative power while minimizing the risk of overfitting in small-sample settings. Evaluation used 5-fold stratified cross-validation with held-out predictions. Results were benchmarked against an equivalent model trained on morphological foundation embeddings.
- **Unsupervised zero-shot stratification**: To evaluate the inherent structure of predicted programs without outcome-specific training, patient-level program scores were reduced via Principal Component Analysis (PCA) and clustered using K-Means (*k* = 2). Association between cluster membership and clinical response (pCR for breast cancer; CA-125 response for ovarian cancer) was tested via Fisher’s exact test [50], reporting odds ratios (OR) with 95% confidence intervals.
- **Differential program analysis**: To characterize the biological differences between responders and non-responders, Mann–Whitney U tests were performed for each of the 40 programs. Multiple testing correction was applied using the Benjamini–Hochberg procedure to control the false discovery rate (FDR).

All model and clustering hyperparameters, as well as preprocessing (including PCA), were fixed a priori and applied identically across therapy-response cohorts. PCA was applied to both SPARC-Map and image-only embeddings to standardize dimensionality before clustering.

#### Multivariable clinical adjustment

For each cancer type with at least 15 events, we pooled all folds and fit three Cox proportional hazards models on the same patient set after listwise deletion of missing clinical covariates and risk scores: (i) clinical-only (age, sex, grade), (ii) SPARC-Risk only, and (iii) combined (age, sex, grade, SPARC-Risk). We report C-index values for each model and extract the hazard ratio, 95% confidence interval, and *P* -value for the SPARC-Risk term from the combined model. A ridge penalty of 0.01 was used for all models.

#### Covariate C-index comparison

For each cancer type (COAD and READ merged as CRC), we computed per-fold C-indices for age, sex, and grade using univariate Cox models, and a multivariable clinical model (age + sex + grade). SPARC-Risk performance was computed as raw concordance of the model risk scores without Cox refitting. We report the mean ± standard deviation across five folds. Sex-specific cancers were excluded from the sex model, and grade was excluded when fewer than 10 non-null entries were available. Only cancer type–fold combinations with at least 10 patients and 5 events were included. A ridge penalty of 0.01 was used throughout.

#### Implementation details

Training utilized PyTorch DDP across 7 GPUs. We used the Adam optimizer (*lr* = 10^−4^) with a higher learning rate for the fusion gate (2 × 10^−3^) and Stochastic Weight Averaging (SWA) starting at epoch 16.

## Acknowledgments

This research was, in part, funded by the Advanced Research Projects Agency for Health (ARPA-H). The views and conclusions contained in this document are those of the authors and should not be interpreted as representing the official policies, either expressed or implied, of the United States Government. We gratefully acknowledge Andrea Bild, the Susan G. Komen Foundation, Dr. Valentina Nardi, the MIT Abdul Latif Jameel Clinic for Machine Learning in Health, and Ignacio Fuentes for their support, discussions, and contributions to this work. We thank Zelda Mariet and Charlie Saillard from Bioptimus for helpful discus-sions, and Bioptimus for making H-Optimus-1 publicly available.

## Code Availability

All code for SPARC-Map and SPARC-Risk is open-source, and model checkpoints are provided at https://github.com/aziz-ayed/SPARC.

## Data Availability

The results published here are in part based upon data generated by the TCGA Research Net-work (https://www.cancer.gov/tcga). The authors acknowledge the National Cancer Institute and the Foundation for the National Institutes of Health, and their critical role in the creation of the free publicly available NLST dataset. Whole-slide image data for the TCGA, NLST, and the Yale HER2+ Breast and Ovarian treatment-response cohorts are available via The Cancer Imaging Archive (TCIA) at https://www.cancerimagingarchive.net. We also acknowledge the use of the SurGen dataset, available at https://www.ebi.ac.uk/ biostudies/bioimages/studies/S-BIAD1285, for evaluating colorectal cancer morphology, and the HEST-1k dataset, available at https://huggingface.co/datasets/MahmoodLab/hest, for spatial transcriptomics evaluation. The authors thank the patients and clinical providers who contributed to the MGH institutional cohort, which is not publicly available due to data use restrictions. Model inference uses only H&E slides at deployment; no paired molecular assays are required for inference.

## Supplementary Figures

**Supplementary Fig. 1.**
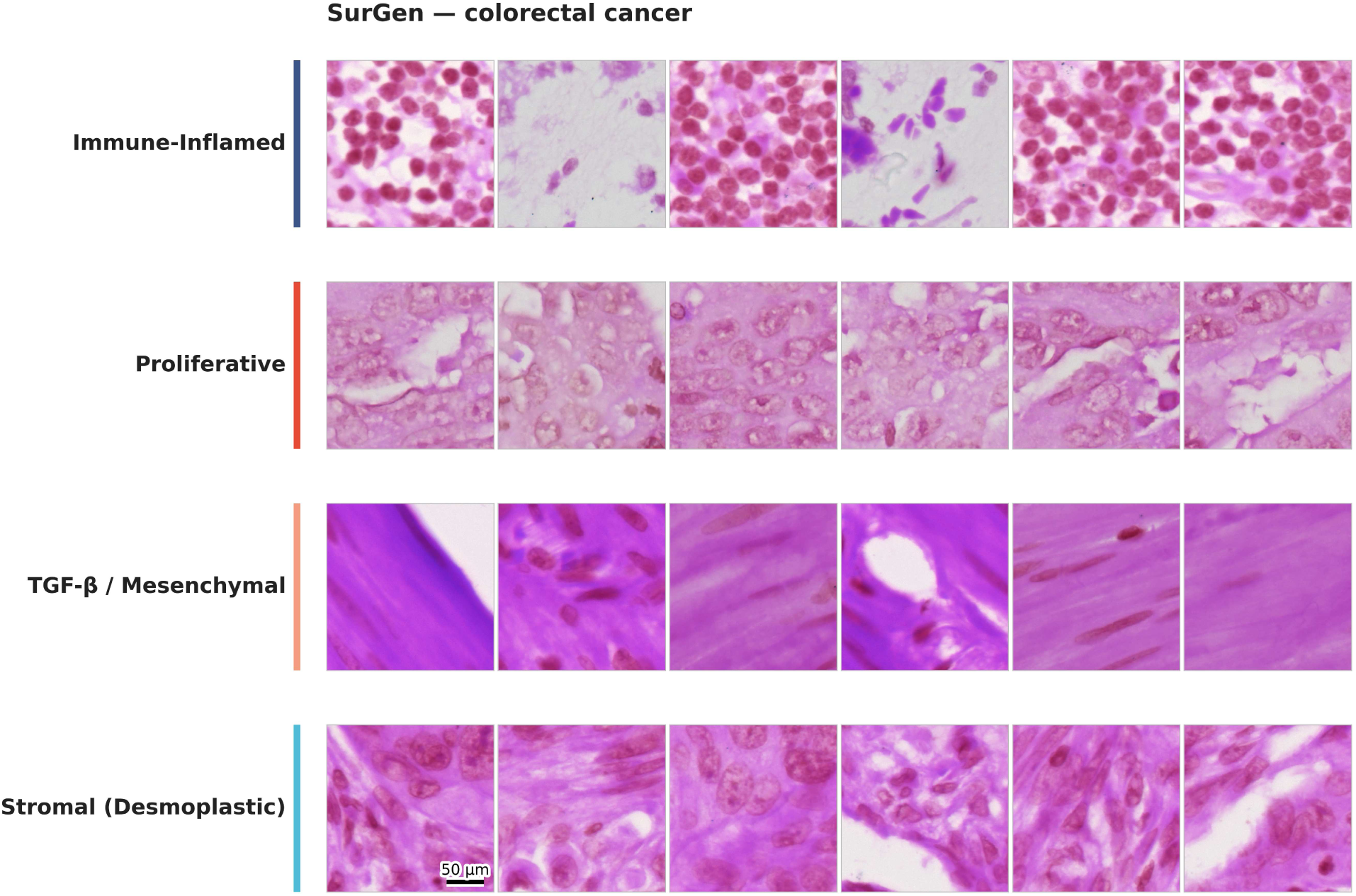
Representative niche morphology in SurGen colorectal cancer. Six representative H&E patches for each niche are shown, drawn from three representative slides per niche. Row colors match the niche palette in Figure 3b. Scale bar: 50 *µ*m.

**Supplementary Fig. 2.**
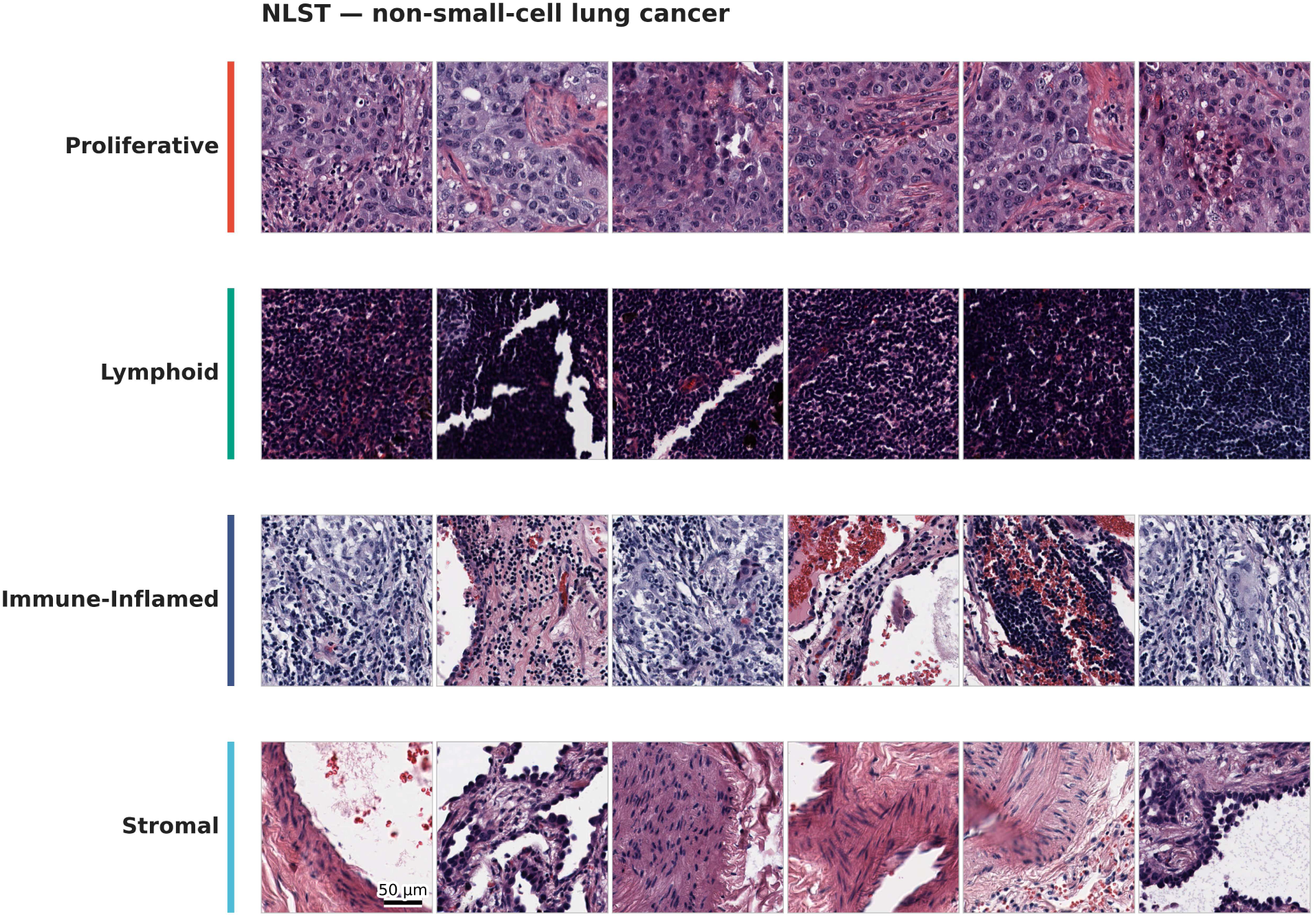
Representative niche morphology in NLST non-small-cell lung cancer. Six representative H&E patches for each niche are shown, drawn from three representative slides per niche. Row colors match the niche palette in Figure 3b. Scale bar: 50 *µ*m.

**Supplementary Fig. 3.**
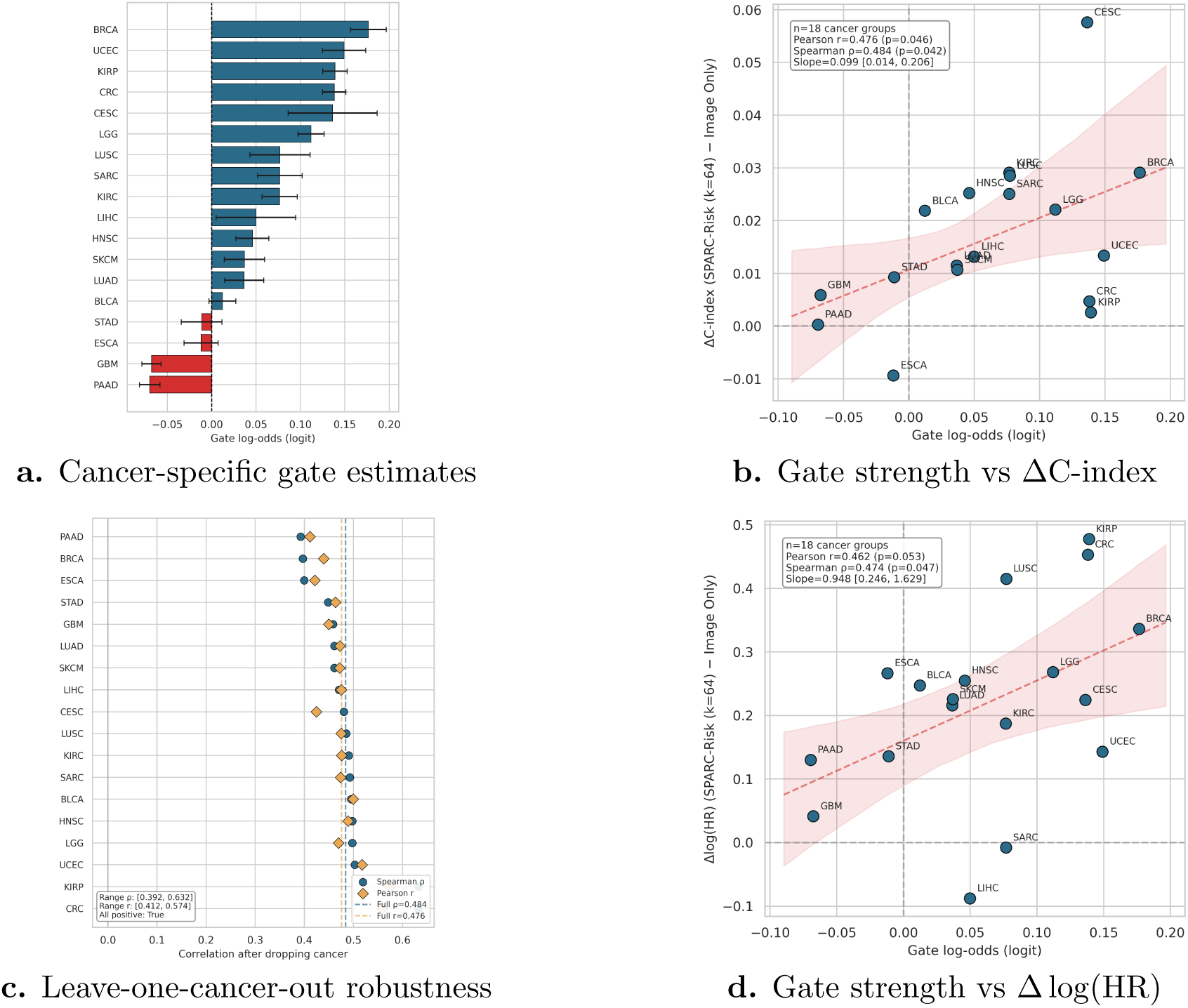
Learned, cancer-specific gating up-weights or down-weights the biological enrichment pathway and tracks prognostic gain. **a.** Learned per-cancer gate log-odds (logit scale) modulate the biological pathway relative to morphology in SPARC-Risk; positive values indicate up-weighting of biological program information, while negative values indicate down-weighting toward morphology-dominant prediction. **b.** Gate strength versus discrimination gain over the image-only baseline, measured as ΔC-index. **c.** Leave-one-cancer-out analysis showing Pearson and Spearman associations remain positive when any single cancer type is removed. **d.** Gate strength versus improvement in risk separation over image-only, quantified as Δ log(HR) between high- and low-risk groups. Together, these analyses show that SPARC-Risk learns cancer-dependent pathway weighting: it up-weights biology where it adds prognostic signal and down-weights it where morphology already captures most of the signal.

**Supplementary Fig. 4.**
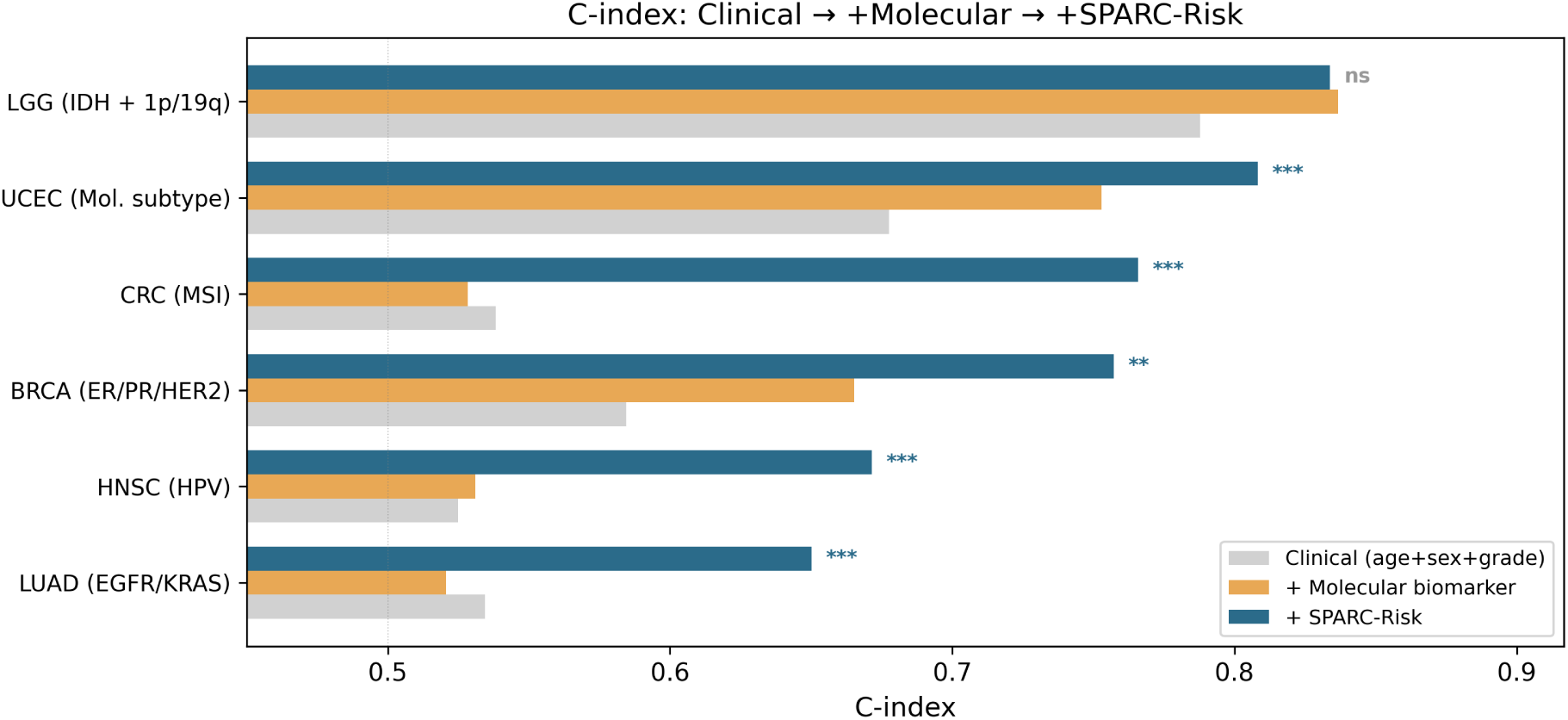
Incremental prognostic value of SPARC-Risk beyond clinical and molecular covariates. Bar plot summary across six molecularly annotated TCGA cohorts (LGG, BRCA, CRC, UCEC, HNSC, LUAD), comparing C-index for clinical-only models (age, sex, grade), clinical + molecular models, and clinical + molecular + SPARC-Risk models. Adding SPARC-Risk to clinical + molecular covariates improved C-index in 5/6 cancers (mean ΔC-index = +0.109), with the largest gains in CRC (+0.238), HNSC (+0.141), and LUAD (+0.129), and a minimal decrement in LGG (−0.003).

**Supplementary Fig. 5.**
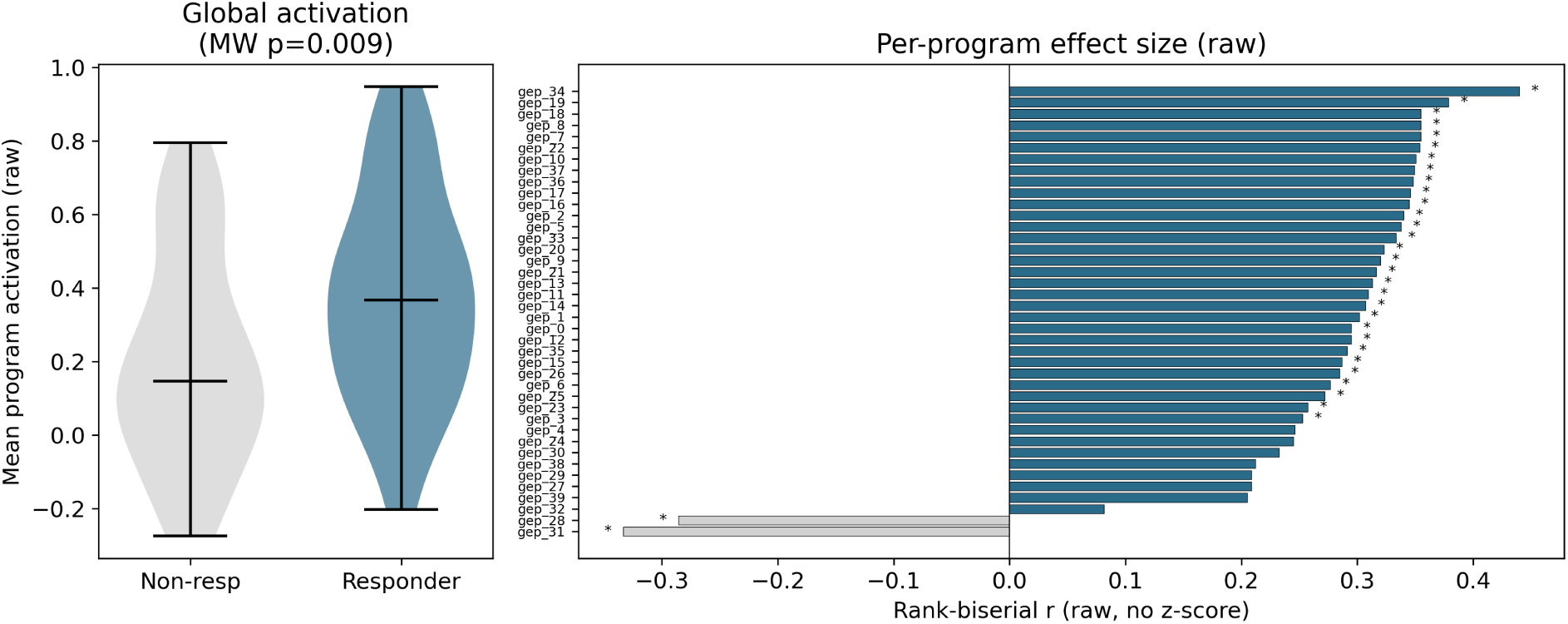
Breast cohort raw-space pan-activation and per-program effect sizes. Left: global activation summary using non-z-scored program scores shows higher mean activation in responders (Mann–Whitney *P* = 0.009). Right: per-program rank-biserial effect sizes in raw space indicate broad upward shifts in responders across 38 of 40 programs, consistent with a pan-active responder state.

**Supplementary Fig. 6.**
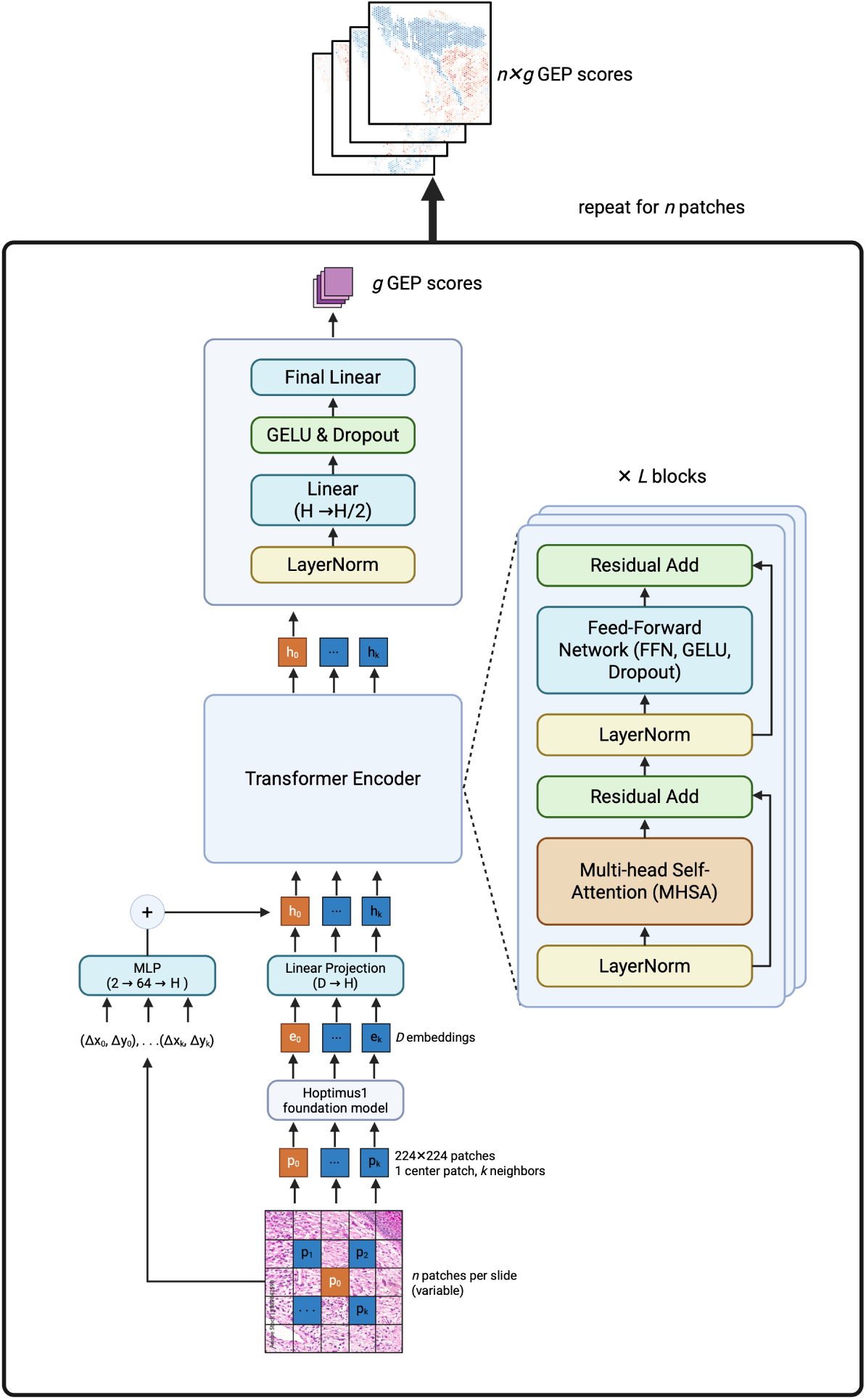
Architecture diagram of SPARC-Map. Predictions are performed per patch. A 224×224 center patch and *k* neighbor patches are passed through the Hoptimus1 foundation model and a linear projection layer to obtain patch-level embeddings. Relative neighbor coordinates are embedded through a linear projection and added to the Hoptimus1 embeddings. Patches are then processed by a transformer encoder with *L* blocks. The output embedding for the center patch is passed through an MLP head to produce final GEP score predictions.

**Supplementary Fig. 7.**
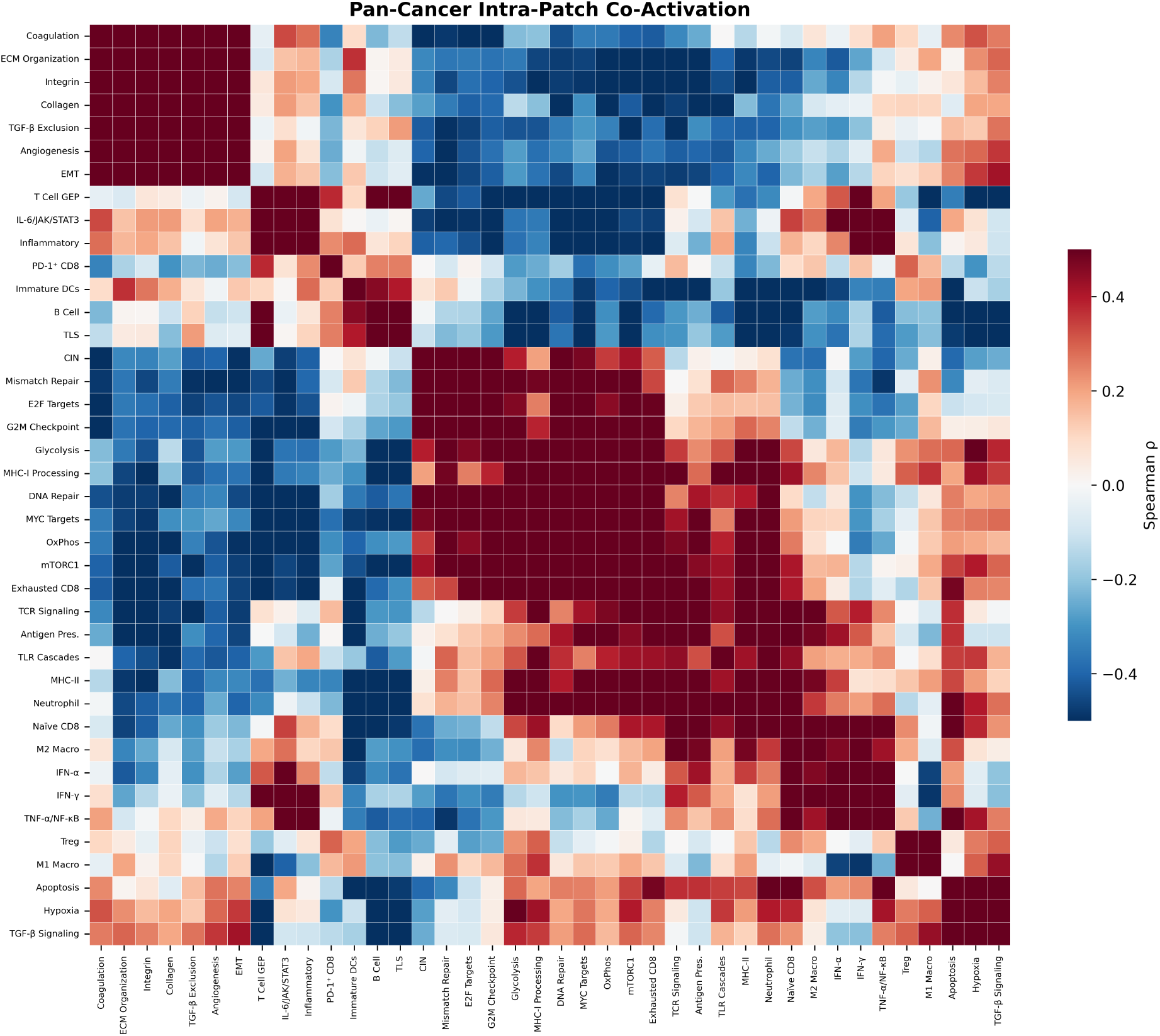
Pan-cancer co-activation matrix. Clustered 40 40 Spear-man correlation heatmap of within-patch program co-activation across 8,796 TCGA slides, computed after per-patch centering. Functional modules (proliferative, stromal, lymphoid, immune signaling) and anti-correlated blocks are visible.

**Supplementary Fig. 8.**
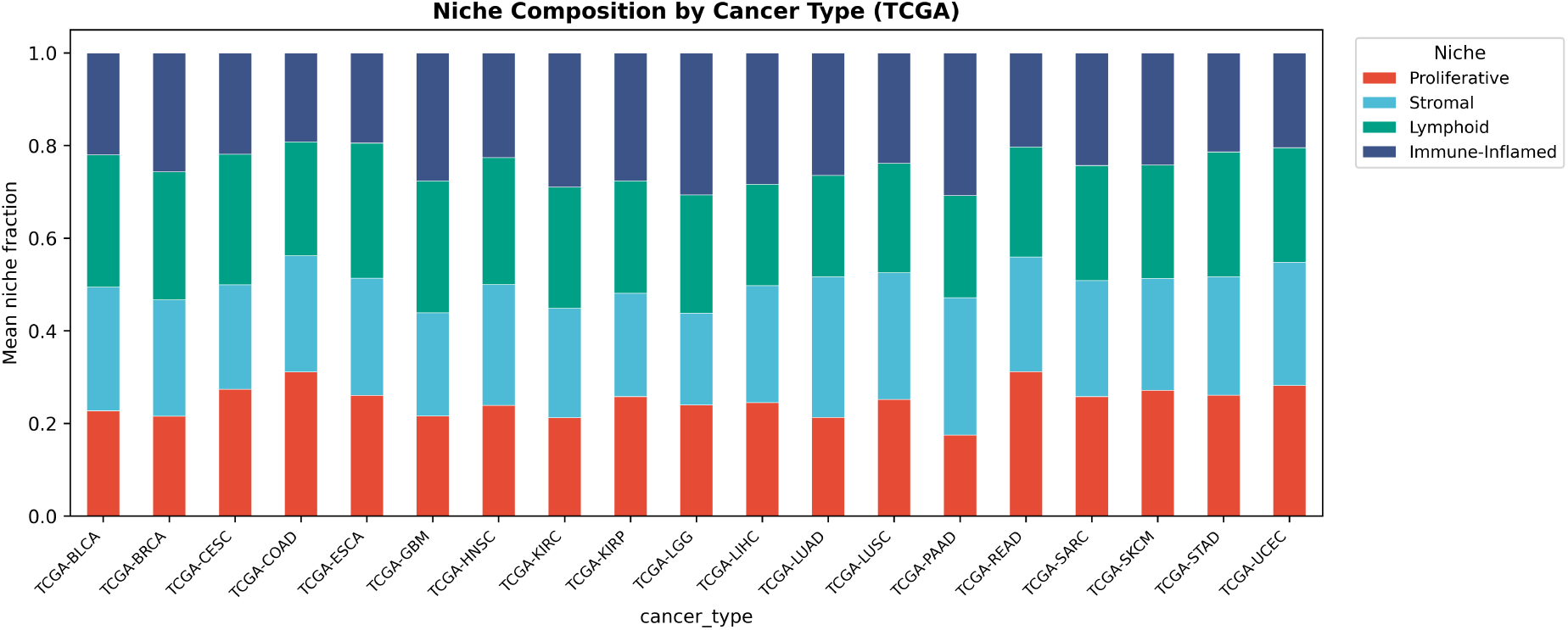
Per-cancer niche composition. Stacked bar chart showing the fraction of each niche (Proliferative, Stromal, Lymphoid, Immune-inflamed) across all 19 TCGA cancer types.

**Supplementary Fig. 9.**
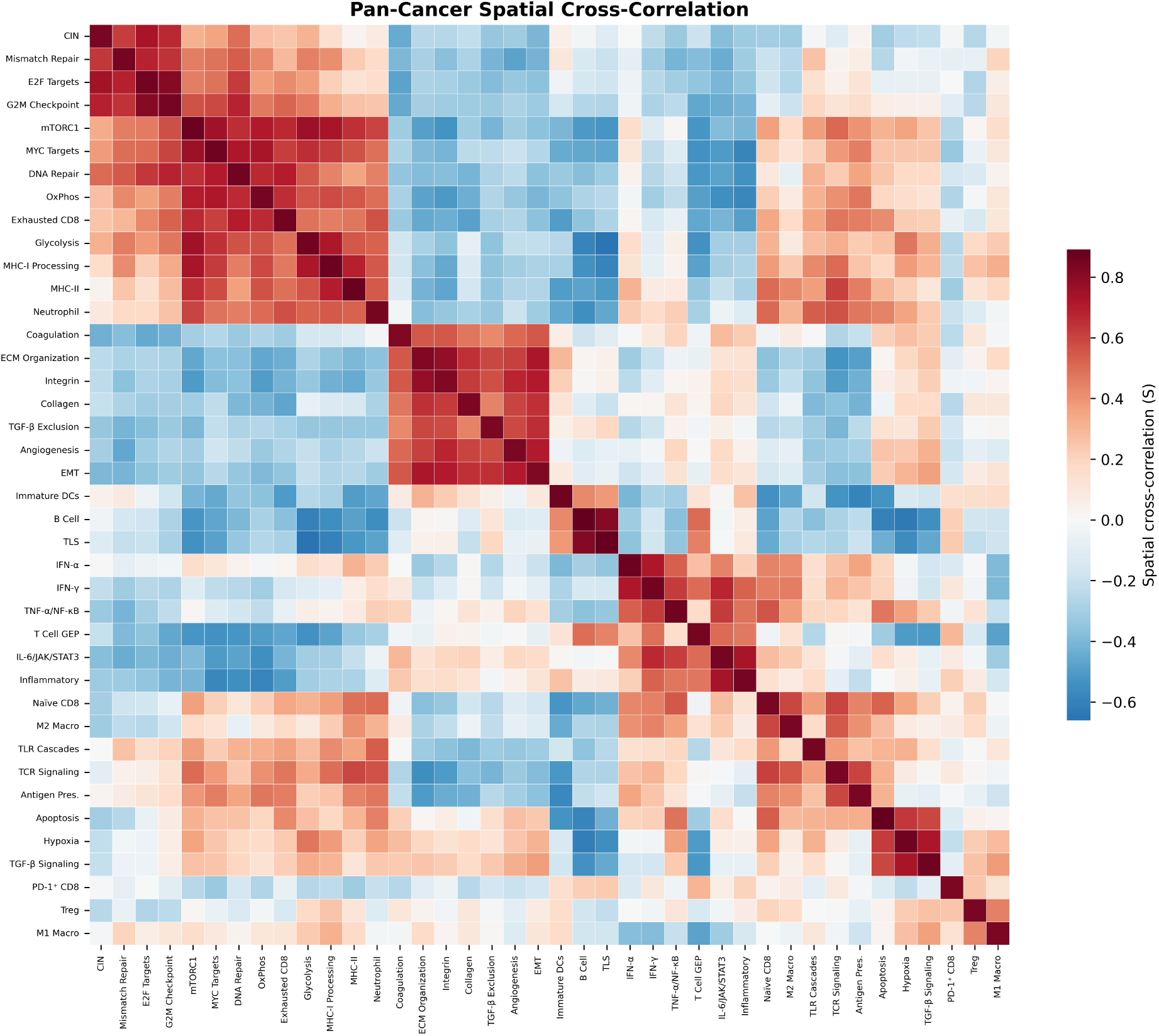
Spatial cross-correlation heatmaps. Full 40 × 40 spatial cross-correlation (*S*) heatmaps for the pan-cancer analysis and selected cancer types, showing cancer-specific variation in spatial program relationships.

**Supplementary Fig. 10.**
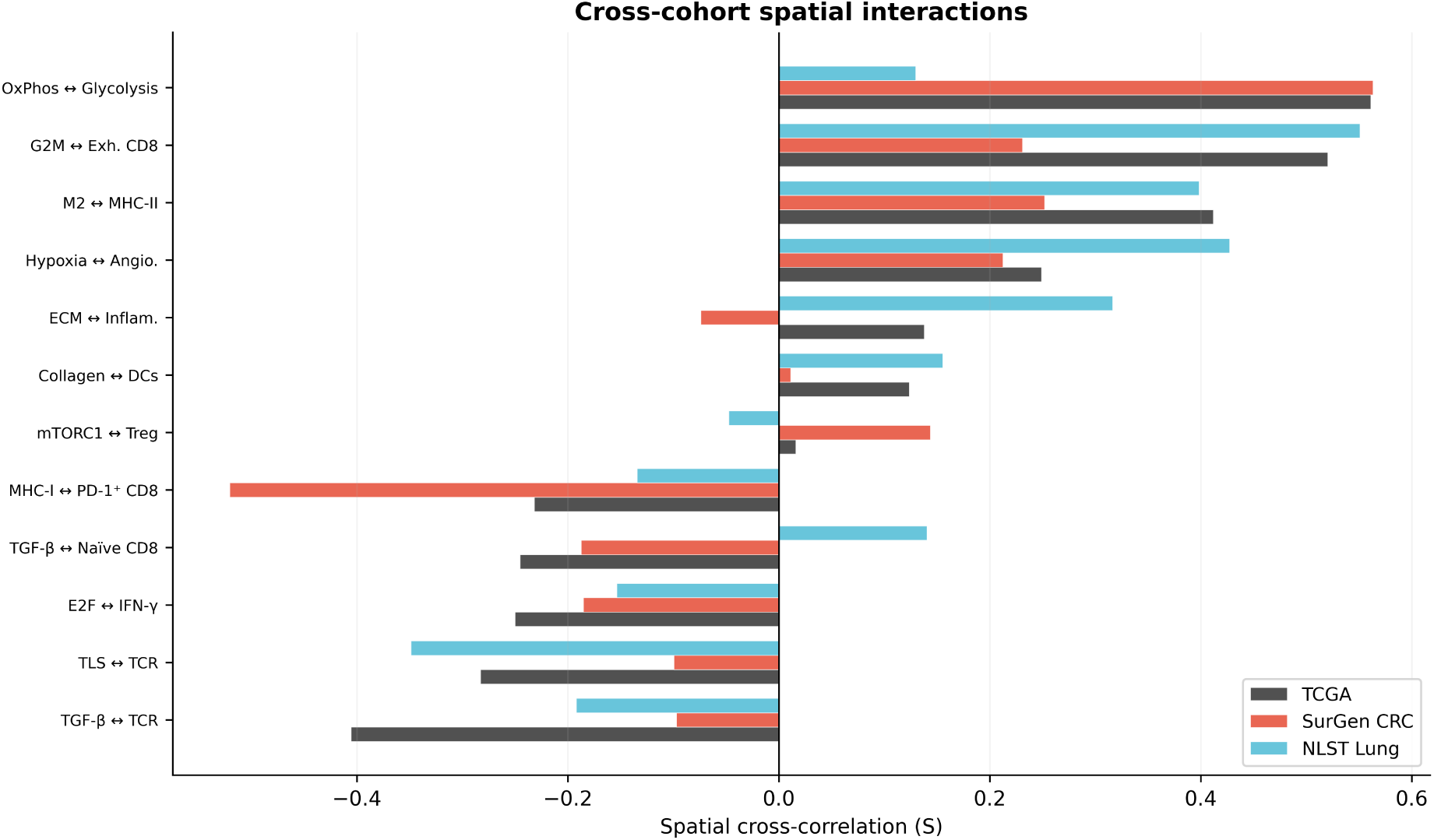
Cross-cohort spatial interaction comparison. Spatial cross-correlation (*S*) values for predefined program pairs compared across TCGA (pan-cancer), SurGen (CRC), and NLST (lung). Conserved spatial relationships (for example, OxPhos Glycolysis co-localization) and tissue-specific patterns are visible.

## Supplementary Tables

**Supplementary Table 1.**
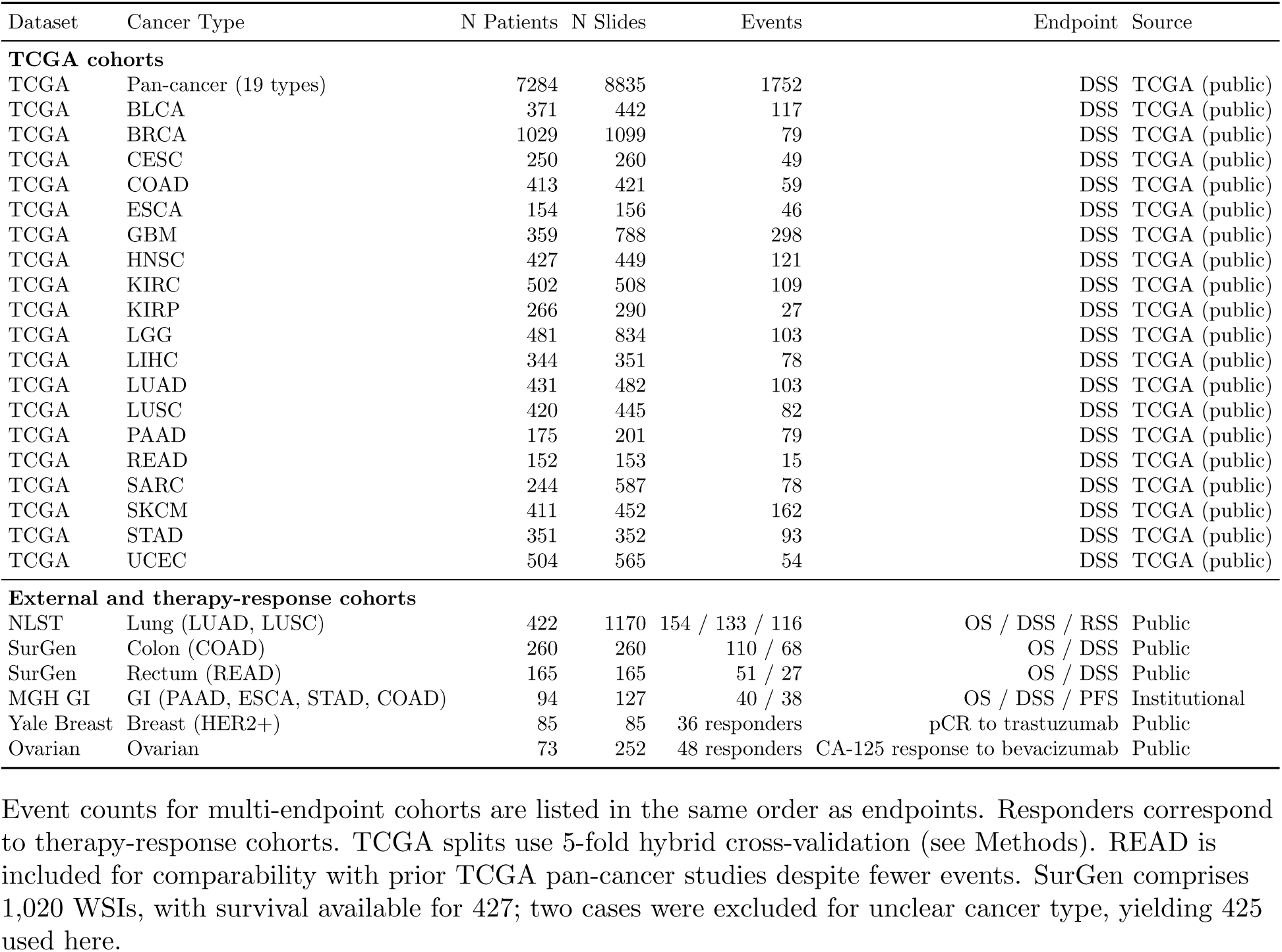
Cohort summary.

| Dataset | Cancer Type | N Patients | N Slides | Events | Endpoint | Source |
| --- | --- | --- | --- | --- | --- | --- |
| **TCGA cohorts** |  |  |  |  |  |  |
| TCGA | Pan-cancer (19 types) | 7284 | 8835 | 1752 | DSS | TCGA (public) |
| TCGA | BLCA | 371 | 442 | 117 | DSS | TCGA (public) |
| TCGA | BRCA | 1029 | 1099 | 79 | DSS | TCGA (public) |
| TCGA | CESC | 250 | 260 | 49 | DSS | TCGA (public) |
| TCGA | COAD | 413 | 421 | 59 | DSS | TCGA (public) |
| TCGA | ESCA | 154 | 156 | 46 | DSS | TCGA (public) |
| TCGA | GBM | 359 | 788 | 298 | DSS | TCGA (public) |
| TCGA | HNSC | 427 | 449 | 121 | DSS | TCGA (public) |
| TCGA | KIRC | 502 | 508 | 109 | DSS | TCGA (public) |
| TCGA | KIRP | 266 | 290 | 27 | DSS | TCGA (public) |
| TCGA | LGG | 481 | 834 | 103 | DSS | TCGA (public) |
| TCGA | LIHC | 344 | 351 | 78 | DSS | TCGA (public) |
| TCGA | LUAD | 431 | 482 | 103 | DSS | TCGA (public) |
| TCGA | LUSC | 420 | 445 | 82 | DSS | TCGA (public) |
| TCGA | PAAD | 175 | 201 | 79 | DSS | TCGA (public) |
| TCGA | READ | 152 | 153 | 15 | DSS | TCGA (public) |
| TCGA | SARC | 244 | 587 | 78 | DSS | TCGA (public) |
| TCGA | SKCM | 411 | 452 | 162 | DSS | TCGA (public) |
| TCGA | STAD | 351 | 352 | 93 | DSS | TCGA (public) |
| TCGA | UCEC | 504 | 565 | 54 | DSS | TCGA (public) |
| **External and therapy-response cohorts** |  |  |  |  |  |  |
| NLST | Lung (LUAD, LUSC) | 422 | 1170 | 154 / 133 / 116 | OS / DSS / RSS | Public |
| SurGen | Colon (COAD) | 260 | 260 | 110 / 68 | OS / DSS | Public |
| SurGen | Rectum (READ) | 165 | 165 | 51 / 27 | OS / DSS | Public |
| MGH GI | GI (PAAD, ESCA, STAD, COAD) | 94 | 127 | 40 / 38 | OS / DSS / PFS | Institutional |
| Yale Breast | Breast (HER2+) | 85 | 85 | 36 responders | pCR to trastuzumab | Public |
| Ovarian | Ovarian | 73 | 252 | 48 responders | CA-125 response to bevacizumab | Public |

Event counts for multi-endpoint cohorts are listed in the same order as endpoints. Responders correspond to therapy-response cohorts. TCGA splits use 5-fold hybrid cross-validation (see Methods). READ is included for comparability with prior TCGA pan-cancer studies despite fewer events. SurGen comprises 1,020 WSIs, with survival available for 427; two cases were excluded for unclear cancer type, yielding 425 used here.

**Supplementary Table 2.**
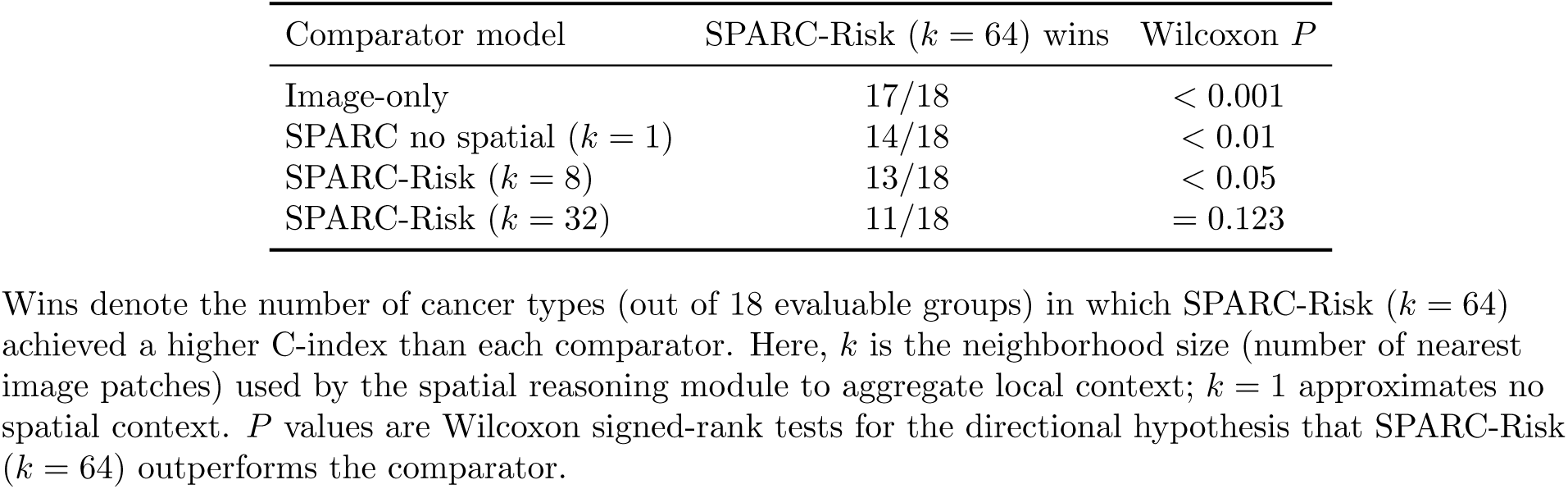
Spatial ablation versus the full SPARC-Risk model (*k* = 64).

**Supplementary Table 3.**
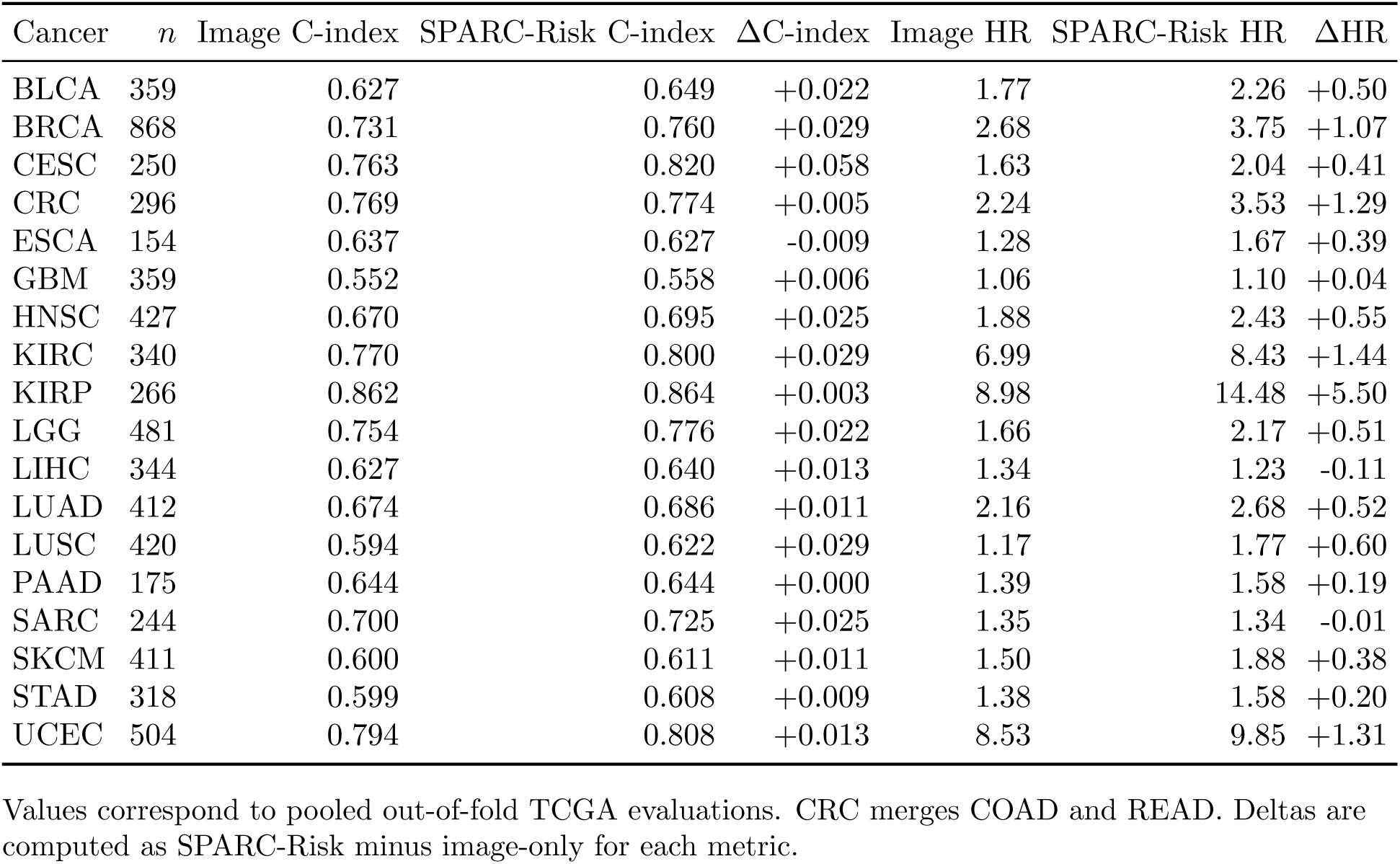
Per-cancer discrimination and risk-separation metrics underlying TCGA pan-cancer results.

**Supplementary Table 4.** Covariate C-index comparison across 5-fold cross-validation.

| Cancer | Age | Sex | Grade | All Clinical | SPARC-Risk |
| --- | --- | --- | --- | --- | --- |
| BLCA | 0.565 ± 0.072 | 0.526 ± 0.015 | 0.537 ± 0.000 | 0.645 ± 0.057 | **0.651 ± 0.031** |
| BRCA | 0.554 ± 0.068 | — | — | 0.554 ± 0.068 | **0.776 ± 0.049** |
| CESC | — | — | — | — | — |
| CRC | 0.690 ± 0.100 | 0.571 ± 0.039 | — | 0.713 ± 0.082 | **0.762 ± 0.087** |
| ESCA | 0.575 ± 0.047 | 0.558 ± 0.052 | 0.641 ± 0.048 | **0.696 ± 0.045** | 0.612 ± 0.078 |
| GBM | 0.608 ± 0.049 | 0.531 ± 0.011 | — | **0.615 ± 0.053** | 0.518 ± 0.019 |
| HNSC | 0.537 ± 0.017 | 0.530 ± 0.035 | 0.563 ± 0.017 | 0.612 ± 0.032 | **0.644 ± 0.040** |
| KIRC | 0.638 ± 0.073 | 0.563 ± 0.057 | 0.787 ± 0.083 | **0.820 ± 0.082** | 0.799 ± 0.105 |
| KIRP | 0.545 ± 0.028 | 0.656 ± 0.091 | — | 0.669 ± 0.108 | **0.950 ± 0.023** |
| LGG | 0.721 ± 0.083 | 0.530 ± 0.039 | 0.681 ± 0.058 | **0.805 ± 0.031** | 0.734 ± 0.078 |
| LIHC | 0.563 ± 0.080 | 0.545 ± 0.014 | 0.594 ± 0.057 | **0.637 ± 0.053** | 0.615 ± 0.153 |
| LUAD | 0.555 ± 0.045 | 0.525 ± 0.044 | — | 0.580 ± 0.023 | **0.684 ± 0.099** |
| LUSC | 0.592 ± 0.061 | 0.552 ± 0.046 | — | **0.620 ± 0.065** | 0.607 ± 0.051 |
| PAAD | 0.567 ± 0.049 | 0.529 ± 0.029 | 0.604 ± 0.089 | 0.630 ± 0.060 | **0.644 ± 0.107** |
| SARC | 0.590 ± 0.030 | 0.566 ± 0.062 | — | **0.634 ± 0.036** | 0.622 ± 0.038 |
| SKCM | 0.599 ± 0.050 | 0.529 ± 0.024 | — | **0.605 ± 0.047** | 0.594 ± 0.069 |
| STAD | 0.545 ± 0.025 | 0.553 ± 0.053 | 0.559 ± 0.047 | **0.616 ± 0.090** | 0.607 ± 0.079 |
| UCEC | 0.589 ± 0.041 | — | 0.678 ± 0.055 | 0.721 ± 0.066 | **0.799 ± 0.068** |
| Avg. | 0.590 | 0.551 | 0.627 | 0.657 | **0.683** |

Best C-index per cancer is shown in bold and the second best is underlined. CRC combines COAD and READ; sex-specific cancers are excluded from the sex model. Values are mean ± s.d. across folds.

**Supplementary Table 5.** Breast trastuzumab cohort clinical stratification and feature-ablation analysis.

| Panel | Comparison | *n* | Response Rate / LOOCV AUC | AUC 95% CI | OR | 95% CI | *P* |
| --- | --- | --- | --- | --- | --- | --- | --- |
| **Panel A. Baseline clinical stratification (Fisher exact)** |  |  |  |  |  |  |  |
| ER status ($\geq 1\%$) | ER- vs ER+ | 22 vs 63 | 40.9% vs 42.9% | — | 0.92 | [0.34, 2.47] | 1.000 |
| HR status (ER+ or PR+ $\geq 1\%$) | HR- vs HR+ | 19 vs 66 | 36.8% vs 43.9% | — | 0.74 | [0.26, 2.13] | 0.6109 |
| **Panel B. Feature ablation** |  |  |  |  |  |  |  |
| Clinical only (ER + PR + HER2 IHC) | — | 85 | 0.451 | [0.337, 0.564] | 1.15 | [0.44, 2.96] | 0.8141 |
| GEP only (40 programs) | — | 85 | 0.588 | [0.467, 0.710] | 2.85 | [1.08, 7.50] | 0.0395* |
| Embeddings only | — | 85 | 0.561 | [0.439, 0.678] | 1.27 | [0.51, 3.12] | 0.6538 |
| Clinical + GEP | — | 85 | 0.623 | [0.502, 0.741] | 3.38 | [1.24, 9.17] | 0.0203* |
| Clinical + Embeddings | — | 85 | 0.614 | [0.495, 0.727] | 2.50 | [1.00, 6.26] | 0.0726 |
| **Panel C. Subgroup rescue within ER+ (Luminal HER2+)** |  |  |  |  |  |  |  |
| GEP clusters | Cluster 0 vs Cluster 1 | 49 vs 14 | 35% vs 71% | — | 0.21* | [0.06, 0.78] | 0.0296 |

In Panel B, AUC values are reported from LOOCV with 95% confidence intervals. ORs are reported from Fisher exact tests with 95% confidence intervals. In Panel C, the OR is reported for Cluster 0 vs Cluster 1; Cluster 1 shows the higher response rate. \* indicates $P < 0.05$ .

**Supplementary Table 6.** Ovarian bevacizumab cohort feature-ablation analysis with baseline CA-125.

| Feature Set | LOOCV AUC | AUC 95% CI | OR | 95% CI | *P* |
| --- | --- | --- | --- | --- | --- |
| CA-125 only | 0.446 | [0.318, 0.575] | 0.48 | [0.16, 1.43] | 0.2059 |
| GEP only (40 programs) | 0.823 | [0.718, 0.906] | 8.07 | [2.71, 24.05] | 0.0001* |
| Embeddings only | 0.541 | [0.410, 0.674] | 1.12 | [0.41, 3.10] | 1.0000 |
| CA-125 + GEP | 0.813 | [0.713, 0.896] | 6.33 | [2.12, 18.96] | 0.0011* |
| CA-125 + Embeddings | 0.614 | [0.477, 0.739] | 1.98 | [0.74, 5.28] | 0.2136 |

AUC values are reported from LOOCV with 95% confidence intervals. ORs are reported from Fisher exact tests with 95% confidence intervals. \* indicates $P < 0.05$ .

**Supplementary Table 7.**
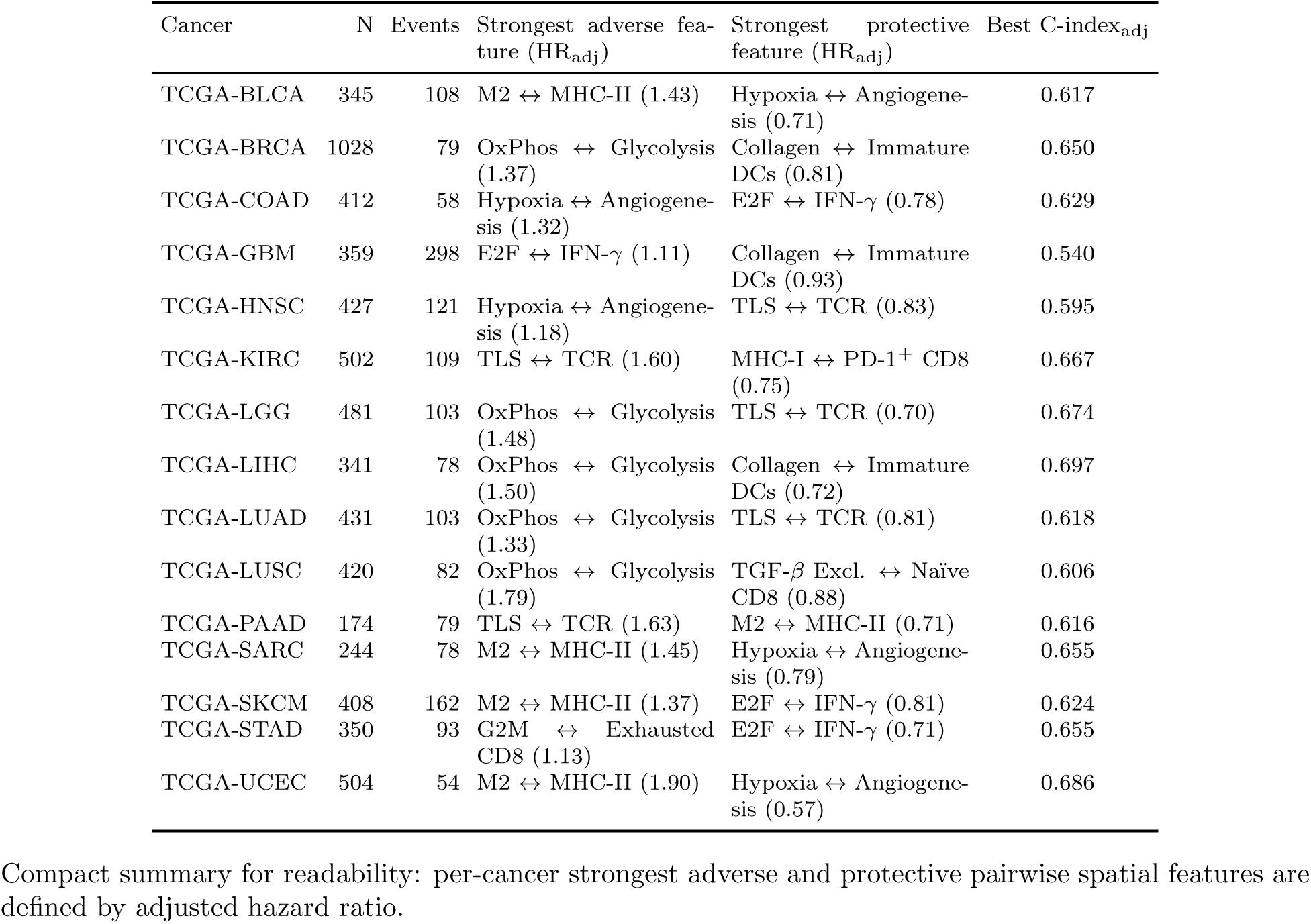
TCGA pan-cancer spatial feature survival summary.

**Supplementary Table 8.** SurGen CRC spatial feature survival summary.

| Subtype | Endpoint | N | Events | Strongest adverse feature (HR) | Strongest protective feature (HR) | Best C-index |
| --- | --- | --- | --- | --- | --- | --- |
| COAD | DSS | 259 | 67 | Stromal (1.94) | Lymphoid (TGF-β exclusion) (0.67) | 0.695 |
| COAD | OS | 259 | 109 | Stromal (1.73) | TGF-β Excl. ↔ TCR (0.72) | 0.661 |
| READ | DSS | 165 | 27 | OxPhos ↔ Glycolysis (3.32) | E2F ↔ IFN-γ (0.47) | 0.733 |
| READ | OS | 165 | 51 | OxPhos ↔ Glycolysis (2.05) | E2F ↔ IFN-γ (0.62) | 0.654 |
| All | DSS | 424 | 94 | Stromal (1.48) | Lymphoid (TGF-β exclusion) (0.66) | 0.622 |
| All | OS | 424 | 160 | OxPhos ↔ Glycolysis (1.32) | TGF-β Excl. ↔ TCR (0.75) | 0.591 |

Compact summary for readability: per subgroup-endpoint strongest adverse and protective spatial features are defined by hazard ratio.

**Supplementary Table 9.**
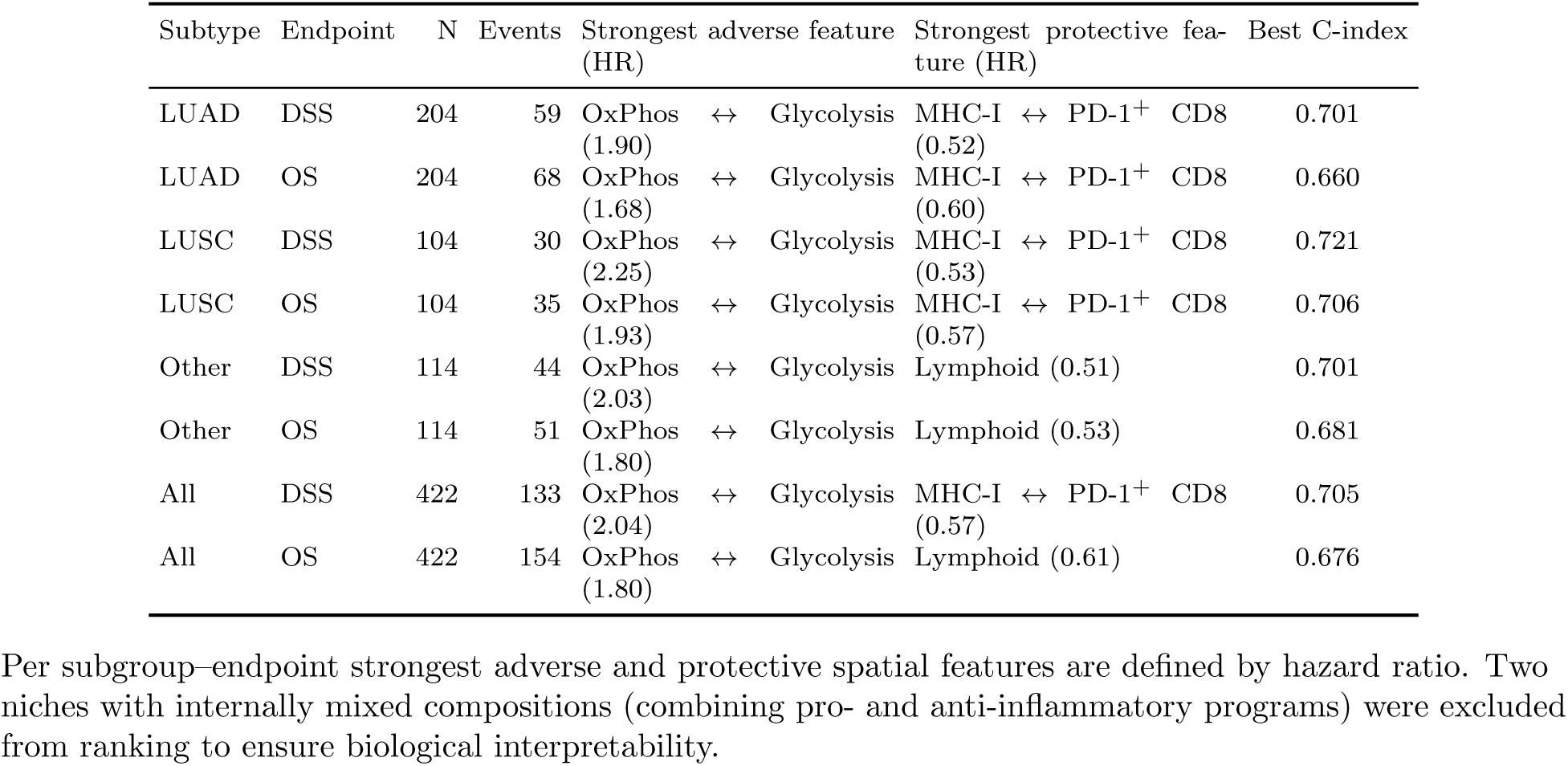
NLST lung spatial feature survival summary.

**Supplementary Table 10.**
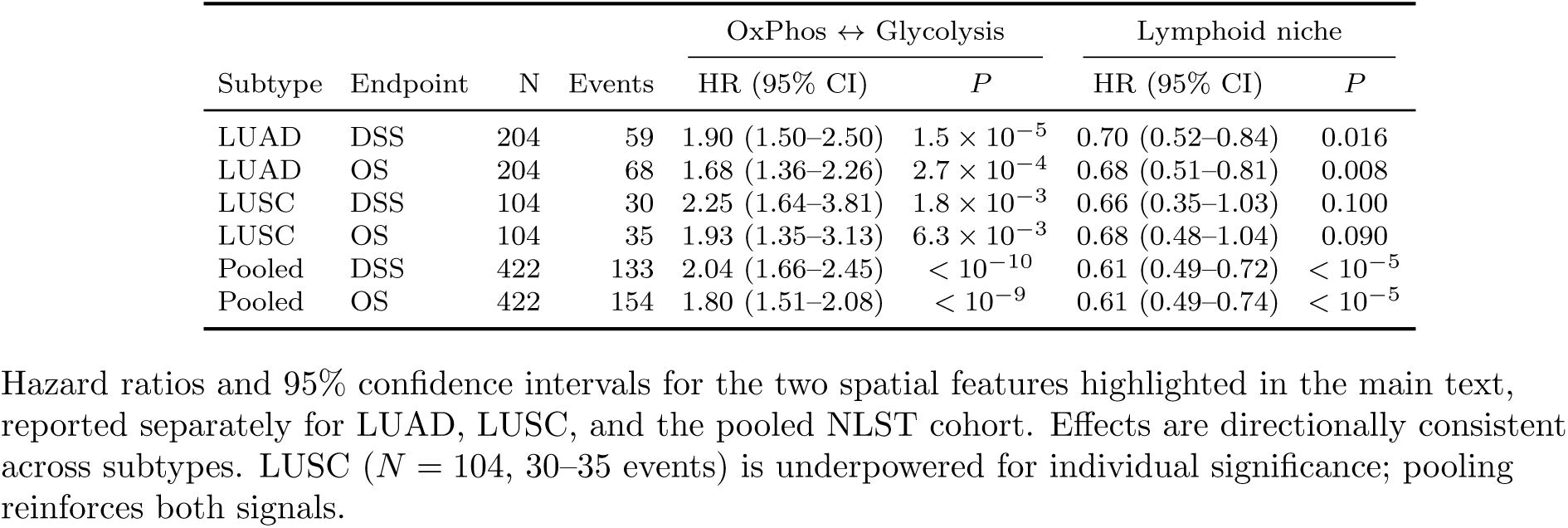
NLST per-subtype survival results for main-text spa-tial features.

| Subtype | Endpoint | N | Events | OxPhos ↔ Glycolysis |  | Lymphoid niche |  |
| --- | --- | --- | --- | --- | --- | --- | --- |
|  |  |  |  | HR (95% CI) | *P* | HR (95% CI) | *P* |
| LUAD | DSS | 204 | 59 | 1.90 (1.50–2.50) | $1.5 \times 10^{-5}$ | 0.70 (0.52–0.84) | 0.016 |
| LUAD | OS | 204 | 68 | 1.68 (1.36–2.26) | $2.7 \times 10^{-4}$ | 0.68 (0.51–0.81) | 0.008 |
| LUSC | DSS | 104 | 30 | 2.25 (1.64–3.81) | $1.8 \times 10^{-3}$ | 0.66 (0.35–1.03) | 0.100 |
| LUSC | OS | 104 | 35 | 1.93 (1.35–3.13) | $6.3 \times 10^{-3}$ | 0.68 (0.48–1.04) | 0.090 |
| Pooled | DSS | 422 | 133 | 2.04 (1.66–2.45) | $< 10^{-10}$ | 0.61 (0.49–0.72) | $< 10^{-5}$ |
| Pooled | OS | 422 | 154 | 1.80 (1.51–2.08) | $< 10^{-9}$ | 0.61 (0.49–0.74) | $< 10^{-5}$ |

Hazard ratios and 95% confidence intervals for the two spatial features highlighted in the main text, reported separately for LUAD, LUSC, and the pooled NLST cohort. Effects are directionally consistent across subtypes. LUSC ( $N = 104$ , 30–35 events) is underpowered for individual significance; pooling reinforces both signals.

**Supplementary Table 11.**
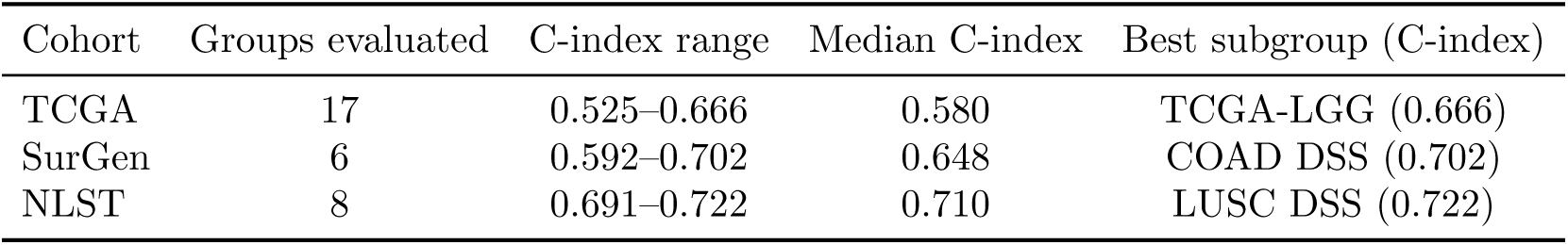
Niche composition multivariable C-index summary.

| Cohort | Groups evaluated | C-index range | Median C-index | Best subgroup (C-index) |
| --- | --- | --- | --- | --- |
| TCGA | 17 | 0.525–0.666 | 0.580 | TCGA-LGG (0.666) |
| SurGen | 6 | 0.592–0.702 | 0.648 | COAD DSS (0.702) |
| NLST | 8 | 0.691–0.722 | 0.710 | LUSC DSS (0.722) |

**Supplementary Table 12.** Clustering diagnostics.

| Cohort | K | Silhouette | Selected | PCA Components | PCA Var. Explained | Stability (cosine) |
| --- | --- | --- | --- | --- | --- | --- |
| TCGA | 4 | 0.1669 | Yes | 8 | 0.909 | > 0.999 |
| TCGA | 5 | 0.1551 | No | 8 | 0.909 | > 0.999 |
| TCGA | 6 | 0.1543 | No | 8 | 0.909 | > 0.999 |
| TCGA | 7 | 0.1511 | No | 8 | 0.909 | > 0.999 |
| TCGA | 8 | 0.1354 | No | 8 | 0.909 | > 0.999 |
| SurGen | 4 | 0.2142 | No | 6 | 0.906 | > 0.999 |
| SurGen | 5 | 0.2204 | Yes | 6 | 0.906 | > 0.999 |
| SurGen | 6 | 0.2039 | No | 6 | 0.906 | > 0.999 |
| SurGen | 7 | 0.1963 | No | 6 | 0.906 | > 0.999 |
| SurGen | 8 | 0.1849 | No | 6 | 0.906 | > 0.999 |
| NLST | 4 | 0.2616 | Yes | 6 | 0.901 | > 0.999 |
| NLST | 5 | 0.2155 | No | 6 | 0.901 | > 0.999 |
| NLST | 6 | 0.2206 | No | 6 | 0.901 | > 0.999 |
| NLST | 7 | 0.1968 | No | 6 | 0.901 | > 0.999 |
| NLST | 8 | 0.1877 | No | 6 | 0.901 | > 0.999 |

